# Moving from shortfall towards adequacy: improving the protein quality of New Zealand vegan diets through optimisation modelling

**DOI:** 10.1101/2025.11.18.25340525

**Authors:** Bi Xue Patricia Soh, Matthieu Vignes, Nick W. Smith, Pamela R. von Hurst, Warren C. McNabb

**Affiliations:** Sustainable Nutrition Initiative, Riddet Institute, Massey University, Palmerston North, New Zealand; School of Mathematical and Computational Sciences, Massey University, Palmerston North, New Zealand; The New Zealand Institute for Bioeconomy Science Ltd (Plant and Food Group), 23 Batchelar Road, Palmerston North 4410, New Zealand; School of Sport Exercise and Nutrition, College of Health, Massey University, Auckland, New Zealand

**Keywords:** Vegan diets, protein quality, amino acids, linear programming, dietary optimisation

## Abstract

**Background:** Poorly planned vegan diets may incur deficiencies in indispensable amino acids (IAAs) and certain micronutrients. Targeted dietary modifications are necessary to improve nutrient adequacy for optimal health.

**Objective:** Optimisation modelling was applied to identify combinations of plant-based foods within an individual’s existing diet to address protein and IAA shortfalls in a sample of New Zealand vegans grouped into three clusters with varied daily dietary patterns.

**Methods:** Shortfalls for protein and IAAs were calculated by comparing daily intakes to individual requirements. An energy-tailored optimisation using linear programming was used: diets with lower energy intake had foods added while those with excess energy had energy-dense and low-protein foods replaced with protein-rich alternatives. The modified diets had to 1) meet protein and IAA shortfalls, 2) respect serving size constraints for added foods, and 3) remain within individual energy boundaries while minimising the weight of food added. Post-optimisation analysis assessed changes in intake of protein, amino acids, dietary fibre and selected micronutrients with results compared across clusters.

**Results:** Protein and IAA shortfalls were more prevalent in cluster 1 (85% of daily diets) compared to clusters 2 (61.1%) and 3 (30.8%). Legumes and pulses contributed most to total protein and lysine with lower energy costs, while nuts and seeds contributed most to methionine and leucine, but with higher energy. Optimisation resolved shortfalls in 93.2% of the daily diets. The remaining 52 diets - mainly from clusters 1 and 2 - could not reach adequacy due to large protein and IAA deficits and limited energy capacity. Post-optimisation micronutrient analysis showed continued risks of shortfalls for calcium, vitamin B12 and iodine.

**Conclusion:** Mathematical optimisation can enhance the protein adequacy of vegan diets while preserving individual acceptability. However, full nutritional adequacy remains challenging in energy-constrained diets with large nutrient deficits.

## 1. Introduction

Foods are not equal in their nutrient provision. For example, the quantities of utilisable indispensable amino acids (IAAs) are generally lower in plant-sourced proteins as compared to animal-sourced proteins (1). This disparity is particularly consequential in vegan diets where the exclusion of all animal-sourced foods means that all protein must come from plant-based (PB) sources. Appropriate quantities and a diversity of PB foods must be consumed to deliver sufficient quantities of these essential nutrients to match human requirements. PB protein sources can be strategically combined to complement one another’s amino acid profiles, hence mitigating shortages of these IAAs in individual foods (2). Food choices are however governed by various factors, including cultural acceptability, individual preferences, and cost of foods, leading to the emergence of varied dietary patterns that differ in diet quality and nutritional profiles.

Devising diets that are both acceptable and nutrient adequate can be challenging in situations where food group diversity is limited, or dietary choices are dominated by energy-dense but nutrient-poor foods. Such combinations restrict the extent to which a diet can be modified to meet nutrient requirements without substantial food substitutions (3). The health benefits of nutrient adequate diets can only be realised if it is accepted and adhered to by the population but defining acceptability constraints is challenging due to the subjectivity associated with individual food preferences and cultural practices (4).

Mathematical optimisation using linear programming (LP) offers an efficient approach to identify the most suitable combinations of available foods that can satisfy nutritional constraints while minimising deviation from the original diet (3, 5, 6). Past modelling techniques have incorporated various strategies to ensure dietary realism, such as setting upper and lower bounds for food intake (3, 4), imposing penalties for deviations from current energy and serving size intakes (4, 7), using each individual’s current dietary intake as the baseline (3, 8) and benchmarking against healthier diets within the same population group (4, 9). LP can be applied in the design of high protein quality vegan diets by measuring the extent to which total protein and utilisable IAA intake can be obtained within the boundaries of individual energy requirements while implicitly accounting for dietary preferences by incorporating some of the above acceptability constraints.

Our prior cluster analysis identified three dietary profiles with varied levels of protein and IAA adequacy with only one cluster close to achieving total protein requirement at the meal level (10). The current or baseline diets of these clusters can be modified through LP to bridge nutritional gaps in protein and IAAs. We hypothesise that the extent of modification necessary will vary across individuals and clusters and influenced by the nutritional quality of the baseline diets – diets that were nearer to sufficiency would require minimal changes in food variety and quantity as compared to those with significant deficits for more than one nutrient. Prolonged deficiencies in amino acids can impair metabolic functions in the body, including protein synthesis and muscle mass maintenance, with certain populations such as the elderly being at higher risks (11). Correcting for these shortfalls through LP in this research provides targeted dietary interventions to improve the overall daily protein intake and protein quality of the vegan cohort, the latter being an aspect often overlooked when considering nutritional adequacies in such populations. While past studies have examined substitutions of animal-sourced proteins with plant-based proteins in varying proportions (12–14), to our knowledge, no studies have applied optimisation methods to systematically improve deficient vegan diets to the point of nutrient adequacy. Given the critical roles of IAAs in metabolic health, and the well-recognised challenges of achieving adequate protein quality while simultaneously balancing energy, fibre and micronutrient needs in the absence of all animal-sourced foods, this study provides a timely and practical contribution. We demonstrate that through planning and use of diverse plant food groups, vegan diets can be configured to meet international protein quality standards. We highlight the feasibility and pathways in diet improvement, offering insights relevant to the growing interest in plant-based diets.

The overall aim of this study was to generate personalised dietary modifications based on each individual’s existing food intake, with the goal of rectifying shortfalls in total protein and IAA, commonly observed in vegan diets. Through the incorporation of familiar foods to the diet, this individualised framework allows diet modifications to remain culturally and behaviourally relevant to each individual (3). A secondary aim is to quantify the deviation in food intake and nutrient contribution required to achieve adequacy by comparing the modified and baseline diets of the population and how these vary across the different clusters. Modified diets should be feasible in implementation, so application of food-based dietary guidelines, such as for serving sizes (6) and energy constraints, serve as boundaries that promote increased likelihood of real-world compliance.

## 2. Methods

### 2.1 Study design and data collection

Vegan dietary data, in the form of four-day food diaries, was obtained from the Vegan Health Research Programme, a cross-sectional study which examined relationships between vegan dietary patterns and nutrition and health outcomes. Data was collected at the Human Nutrition Research Unit at Massey University, Auckland from 2022 to 2023. Ethical approval was granted by the Health and Disability Ethics Committee (HDEC 2022 EXP 12312), and written informed consent was obtained from all participants prior to data collection. As part of the inclusion criteria, only healthy adults above the age of 18 who were not pregnant or breastfeeding and who had been following a vegan diet for at least two years were recruited.

Food items listed in the food diaries were processed with the FoodWorks Professional nutrient analysis software (Xyris, Australia 2022) and matched to appropriate foods found in FOODfiles, the NZ food composition database (15). This provides the nutrient composition, such as total protein, dietary fibre, vitamins, and minerals for each food item. Amino acid (AA) composition is not currently available in FOODfiles. Hence, to quantify the IAA content in NZ foods, each food item was matched to its most similar counterpart from the food and nutrient composition database of the United States Department of Agriculture (USDA) (16) and normalised to the protein content of NZ foods. To indicate the proportion of nutrient that is digestible, protein and IAA compositions were adjusted for digestibility using available True Ileal Digestibility (TID) values (17–20). A full description of steps in data collection and processing can be found in our earlier study (21). We had previously characterised three distinct dietary patterns among this vegan cohort using techniques in time series clustering, which were used here to subdivide the data used in the optimisation (10). Food diaries from 193 individuals, each contributing at least three days of dietary data were used as the baseline dataset for dietary optimisation. After excluding days with missing consumption or time data, a total of 766 daily diets were included to quantify nutrient shortfalls relative to individual requirements

### 2.2 Calculation of individual nutrient requirements and shortfalls

Individual daily protein requirements were calculated based on sex, age and body weight, guided by the Nutrient Reference Values (NRV) for Australia and New Zealand (22). The Estimated Average Requirement (EAR) values were used for this computation. This was 0.68 g/kg of body weight (BW) for males and 0.60 g/kg for females of between 19 to 70 years of age. The EAR was 0.86 g/kg for males and 0.75 g/kg for females above 70 years of age. IAAs with currently available TID values (histidine, leucine, lysine, cystine, methionine, sulphur-amino acids (SAAs), threonine and tryptophan) were included in this study. Individual daily IAA requirements were calculated based on individual body weight, using the reference values from the Food and Agricultural Organisation (FAO) (20). The requirement values for adults 18 years and above are listed in **Table 1**.

**Table 1.**
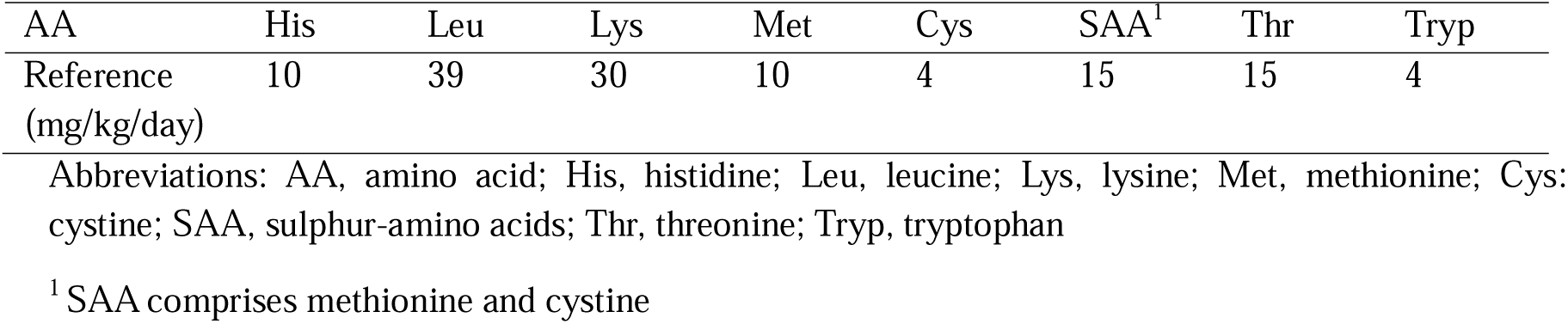
Daily IAA requirement values in mg per kg of body weight per day.

The minimum daily energy requirement in megajoules (MJ) for each individual was calculated by accounting for the sex, age, body weight and height, as well as the physical activity level (PAL) (22). The International Physical Activity Questionnaire (IPAQ) was used to compute the daily PAL (23). Briefly, IPAQ records the number of metabolic equivalent task (MET) minutes, which represents a multiple of the estimated resting energy expenditure. For example, 1 MET indicates energy expended at rest. 3.3 METS, 4 METS and 8 METS represent energy expended during walking, moderate physical activity (PA) and vigorous PA respectively (23). The PAL was then calculated by adding 1 MET to the sum of the average MET values for each physical activity, weighted by the average number of minutes per week spent on that activity (24, 25), as shown:

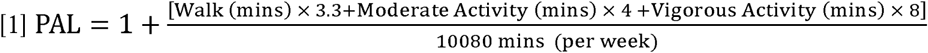

The Estimated Energy Requirement (EER) value was then assigned to each individual, based on the sex, body weight, height, age, and the calculated PAL(22). While an upper limit to dietary energy intake has not been established (22), we applied a 20% increase above the EER as a pragmatic upper boundary. This allowance provided the model with greater flexibility to choose wider range of foods for individuals who met or exceeded their EER but had significant shortfalls for protein and/or IAAs. Shortfalls for protein, IAA and energy (for individuals with daily intake < minimum requirement for EER (EER_min_)) were tabulated for each day, by deducting the total daily intake of each nutrient from the daily requirement. **Figure 1** provides the overall methodology, highlighting the processes in the database preparation and subsequent diet optimisation.

**Figure 1.**
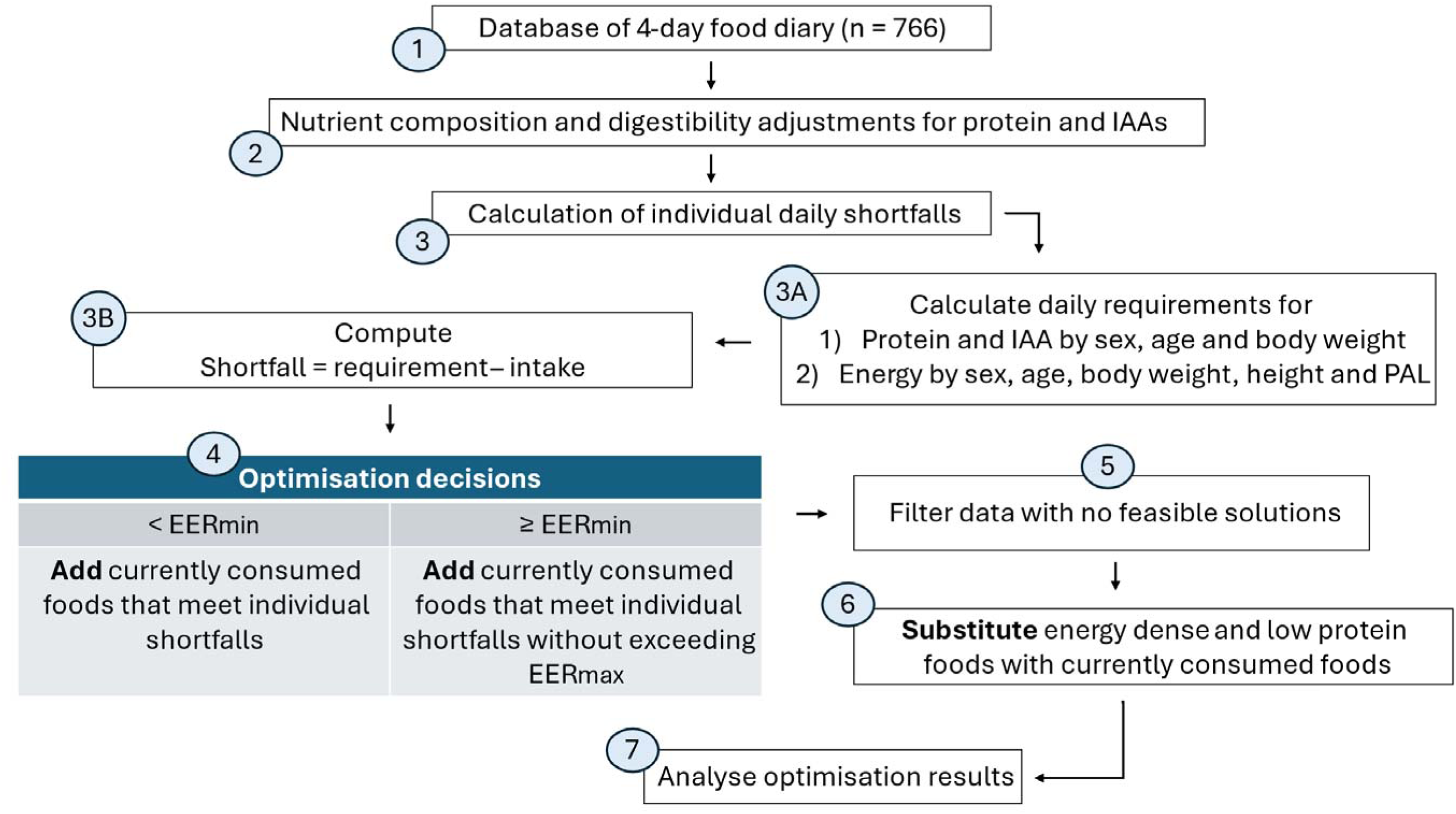
Methodology of LP optimisation; abbreviations: IAA, indispensable amino acids; PAL, physical activity level, EER_min_, minimum estimated energy requirement per day, EER_max_, maximum estimated energy requirement per day. Numbers indicate the steps taken in the optimisation process. (1) Dietary data was in the form of 766 food diaries, recorded by a cohort of 193 NZ vegans. (2) Each food item was matched to its nutrient composition for protein, IAA, energy, dietary fibre and micronutrients in NZ and USDA food composition tables. Total protein and IAAs were adjusted for digestibility. (3) Shortfalls were calculated for each of the 766 daily diets by first, (A) calculating the daily requirements for energy, protein and IAA, with demographic and anthropometric data of each vegan and (B), computing the daily shortfalls by subtracting the baseline daily intake of each nutrient from the daily requirement. (4) LP optimisation methods depended on energy intake of the daily diets. Should energy intake be below (<) EER_min_, additional foods could be added from the individual’s diet to meet protein, IAA and energy shortfalls. Should energy intake be meeting or above (≥) EER_min_, then added foods must meet protein and IAA shortfalls without exceeding EER_max_ (energy is not a shortfall in these cases). (5) After optimisation is completed for all clusters, cases which had no feasible solutions were filtered and combined with cases where added foods had resulted in daily energy intake above EER_max._ (6) For these cases, a substitution strategy was used, to remove foods with highest energy-to-protein ratios to provide energy headroom, so that foods could be added to meet protein and IAA shortfalls. (7) All cases with feasible solutions from step 4 and step 6 were analysed for nutrient intake after optimisation.

### 2.3 Diet optimisation using linear programming (LP)

One of the constraints of our LP model was that foods added should fulfil any protein and IAA shortfalls. Hence, for each nutrient, *i* (where *i* may be protein or one of the IAAs), the quantity, *x_j_* (in grams) of all foods *j* in the optimised diet, must together supply at least the daily requirement, *R*_i_. Formally:

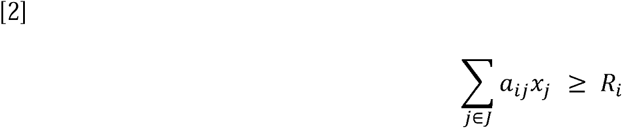

for each nutrient *i*, where:

*a_ij_* is the quantity of nutrient *i* per gram of food *j*, *R_i_* is the daily nutrient requirement for nutrient *i*.

*J* represents the set of all possible food items in the optimised diets

Foods in the optimised diet must result in an energy contribution that is within the lower (EER_min_) and upper (EER_max_) energy boundary for that individual. This is expressed as:

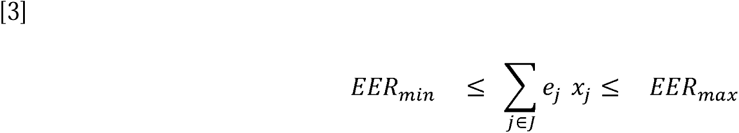

*e_j_* represents the energy (MJ) per gram of food selected, *x_j_*

As described in Figure 1, the optimisation decision is dependent on the daily energy intake (step 4). In all cases, the model is constrained to not exceed EER_max_. If the daily diet is not meeting the EER_min_, then energy is taken as a shortfall and the model chooses foods that will cover the shortfalls for protein, IAA and energy.

The model may not return feasible solutions in every case (step 5). For example, if the current energy intake is close to EER_max_ but significant deficits exist for protein and IAA, it may not be possible to add foods that fill the protein deficit while respecting the energy constraint. A substitution strategy was then utilised to remove the most energy dense, but protein-poor foods from the diet, to accommodate the addition of foods that can fill nutrient gaps (step 6). To do so, an energy-to-protein ratio was calculated for each food in these cases. The first substitution strategy involved removing 25% (by mass) of the original foods that had the highest energy-to-protein ratio, recalculating the new shortfalls and reconducting the LP model to add substitutions. Cases which still had no feasible solutions were then identified. The top 50% of the original foods that had the highest energy-to-protein ratio was then removed, and the substitution process was performed again. Any further cases without feasible solutions were not further addressed, since removal of >50% of food in the baseline diet was seen as too great a deviation to be acceptable to a consumer. Additionally, further removal may not yield significant improvements in terms of energy alignment, nutrient contributions and food diversity (e.g., model may end up choosing very high servings of nuts and seeds).

An acceptability constraint was implicitly introduced through the proxy of adding foods already in an individual’s diet over foods from outside their existing diet. Food items that were already part of the daily diet of each individual were available for selection by the LP model, except for certain foods, which were intentionally omitted at this point. Brazil nuts were removed due to their potential in exceeding selenium upper limits and the narrow window between selenium deficiency, adequacy and toxicity (26); seaweed, which has wide range of iodine content depending on species (0.02 to 428 µg per gram of food in our inventory), could exceed the upper limit of iodine intake (27); and, herbs and spices were not considered as feasible foods to address protein shortfalls in the diet.

The objective function of the LP model was to minimise the total weight of food added to the diet to limit the addition of large volumes of food that cannot be administered in realistic diets. Serving constraints were also set for all food groups to prevent excessive addition of each food item, as guided by the NZ Eating and Activity Guidelines (28). These are listed in **Table 2**.

**Table 2.** Weight of food per serving size for each food group.

| Food group | Maximum weight (g) per serving |
| --- | --- |
| Fruit | 150 |
| Grains and pasta | 120 |
| Legumes and pulses | 150 |
| Nuts and seeds | 30 |
| Potatoes, kumara and taro | 75 |
| Sugar and sweets | 30 |
| Vegetables | 75 |
| Yeast and condiments | 30 |

The serving constraint for spirulina was capped at 3g, according to recommended guidelines from the literature (29, 30). The LP optimisation was constrained to adding up to one serving for each unique food item (but not food group). For example, a maximum of 30 grams of sesame seeds and 30 grams of cashew nuts could be added to address shortfalls. These are different food items, but belong to the same food group, “nuts and seeds”. The characteristics of the optimisation model are described in **Table 3**. Optimisation was conducted separately for each cluster in the vegan cohort and the solutions were compared across clusters. The LP optimisation was completed using R studio (version 4.3.1) with the lp solver package.

**Table 3.** Characteristics of optimisation model.

| Constraints | Objective function | Decision variable | Solution |
| --- | --- | --- | --- |
| 1. Meet protein and IAA shortfalls | Minimise weight of food added | Food items that exist in any day of each individual’s diet | Weight, energy and nutrient contribution of food items added |
| 2. Serving size per food item |  |  |  |
| 3. Optimised diet does not exceed $EER_{max}$ | | | |

As a sensitivity analysis, energy constraints were increased from 20% above EER_min_ to 50% and 100%. Upper limits of each IAA were also introduced to the model at 24% higher than the individual IAA requirements (in mg per kg of body weight) (20). In these scenarios, the number of feasible solutions were identified and impact on optimisation results were assessed. Owing to the importance of some plant protein isolates, such as soy and pea in improving protein quality (31), we also assessed the impact when these foods were removed from the inventory.

### 2.4. Data Analysis: Post-optimisation assessment of total protein and IAA

Results were analysed (Figure 1, step 7) and compared across clusters 1 to 3. The mean weight in grams of added foods for each food group was tabulated. The mean contributions to energy, protein and IAA from each added food group were computed as a percentage of the minimum daily requirement. To obtain the final nutrient intake after optimisation, the quantity of nutrients from foods introduced by optimisation was added to the baseline nutrient intake. For cases where foods had to be substituted (Figure 1, step 6), the diets after the removal of energy-dense foods were used as the new baseline diets for this computation.

Original diets (dietary data in step 1 of Figure 1) and the final optimised diets of all individuals were then compared to determine how energy, protein and IAA contribution per food group changed after the diet modification. Total nutrient intake in the baseline and final diet was also computed as a percentage of the minimum daily requirement. The final protein and IAA intake for all optimised diets were computed with respect to individual body weight (kg) and compared to reference values. For total protein, final intake was compared to the EAR and the Acceptable Macronutrient Distribution Range (AMDR), which is between 10 to 35% of caloric intake (22, 32). For IAAs, the final intake was compared to values in Table 1. Risks of exceeding the upper limit intakes for IAAs were evaluated by comparing quantities against the currently available values of Tolerable Upper Intake Level (UL), No-observed-adverse-effect-level (NOAEL) or Lowest-observed-adverse-effect level (LOAEL) (33) (**Table 4**). For the purposes of this study where we aim to address deficiencies in daily protein and IAA intake, only daily diets with at least one shortfall either in protein or any IAA was optimised and analysed.

**Table 4.**
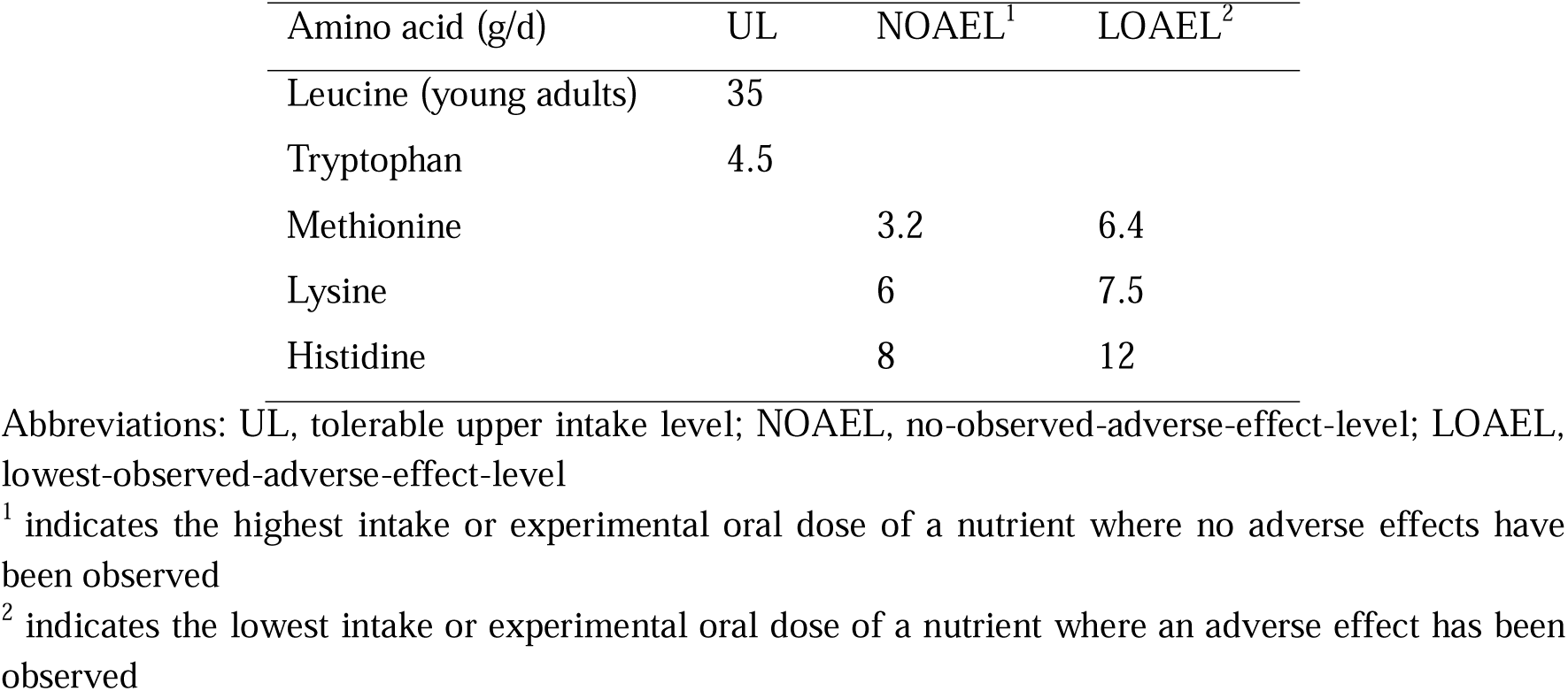
Upper limits of IAAs as adapted from Elango (2023) (33)

### 2.5. Data Analysis: Post-optimisation assessment of changes in other nutrient levels

Finally, dietary fibre, alpha-linolenic acid (ALA) (the most prominent long chain fatty acid from plant-sourced foods) and selected micronutrient quantities of the optimised diets were computed to assess how these nutrients change from baseline. Vitamin B12, calcium, iodine, iron and zinc were the micronutrients focused on in this study because of increased risks of deficiencies in these nutrients as a consequence of vegan diet consumption (34–36). Sodium intake was also analysed against reference values to check if modelled diets had inadvertently added excess sodium. The EAR or adequate intake (AI), when EAR is not available, was used as the minimum daily requirement (22). The UL per day was also provided when applicable (22). This information is provided in **Table S1.** Note that these values were not used as constraints in the optimisation; they are included as further contextual information about the quality of the optimised diets.

## 3. Results

### 3.1 Optimisation results with targeted food additions and substitutions

The number of daily diets with at least one shortfall in cluster 1 was 250, cluster 2 was 215 and cluster 3 was 37. Cluster 1 had the greatest number of days with shortfalls for protein and all IAAs (10% of all days), followed by cluster 2 (4%), and no daily diets in cluster 3 had shortfalls for protein and all IAAs. Lysine, leucine and methionine had the largest average deficits as compared to daily requirement.

The first round of LP optimisation arrived at feasible solutions for 303 daily diets out of 502. For the remaining 199 daily diets, 99 had insufficient residual energy capacity (mean of 1.07 MJ) to accommodate inclusion of additional foods to meet nutrient shortfalls while the rest (n = 100) had exceeded the EER_max._ To resolve shortfalls here, it was necessary to first remove some foods from the baseline diets.

Hence, food substitution was conducted for these 199 cases. When the top 25% of foods with the highest energy-to-protein ratios were removed before optimisation, feasible solutions were obtained for 118 of the 199 cases. Before optimisation, these diets had a mean energy of 3.09 MJ below the EER_max_ and relatively small nutrient shortfalls, mainly in the IAAs. Contrastingly, for the remaining 81 unsolved cases, these diets had a mean energy of 2.14 MJ below the EER_max_ but larger shortfalls, especially for protein. For these 81 cases, further food substitutions were required. The top 50% of foods with the highest energy-to-protein ratios were removed to provide even higher residual energy capacity (mean 4.20 MJ) for new food additions. After this substitution, the LP model was able to find feasible solutions from currently consumed foods for 29 of these cases but 52 cases had no feasible solutions.

Hence, optimisation using addition or substitution methods with currently consumed foods within each individual’s diet obtained solutions for 93.2 % of daily vegan diets which had at least one shortfall at baseline. This was 219 cases in cluster 1, 195 in cluster 2 and 36 in cluster 3. Of the cases where no feasible solutions could be obtained from the current diet (n = 52), cluster 1 diets formed the largest proportion at 60%, followed by cluster 2 (38%).

The mean mass of total food added per person per day was highest for clusters 1 and 2, at 44.9 g (sd = 26.4) and 43.7 g (sd = 27.3) respectively, while the mean mass added for cluster 3 was 39.3 g (sd = 25.8). The mean number of unique foods added for each individual was again higher in clusters 1 and 2, at 3.47 and 3.24 items respectively, while for cluster 3, 2.80 items were added. The most frequently added food items were from the food groups “legumes and pulses” and “nuts and seeds”, with peanut, pumpkin seed, almond, plant-based meat alternatives (PBMAs), tofu and pea protein isolate the most commonly added foods in all clusters.

**Table 5** presents the mean weight and nutrient contribution as a percentage of daily requirements for all added food items for cases in which LP optimisation could find solutions. Food items were categorised to their respective food groups. Large standard deviations (sd) indicated large variation in the quantities of food items added across daily diets. In all the clusters, legumes and pulses contributed most to total protein and IAAs such as threonine, lysine and histidine for the smallest proportion of energy. While nuts and seeds contributed higher quantities of energy per gram of added food, the contribution towards total protein, tryptophan, methionine, leucine and SAA was higher than legumes and pulses. We note the value of yeast-containing food items to nutrient contribution such as lysine and that these foods could be added to the diet with small increases in energy to the diet. The contribution of plant-based alternatives (PBAs) to energy and protein was approximately 10-15% for total protein and IAAs and <10% for energy across all clusters. Almost all PBMAs included pea and soy isolates in the ingredients. These isolates belong to the “legumes and pulses” food group. As fruits were only added in a few cases (two in cluster 1 and four in cluster 2), the average quantity added was inflated and driven by large portions (up to 150 g per fruit can be added) in few individuals. Fruit, as well as sugar and sweets were only added to individuals who had an energy shortfall. Given its relatively high energy content, fruit may have been selected by the LP model as a suitable option from the baseline diet to close the energy gap and a practical fallback in cases where the serving size limits for other food groups - nuts and seeds, legumes and pulses - have reached the maximum serving limit. The mean number of serves added per individual was similar across all clusters and averaged less than 1 serving of each food item per person.

**Table 5.** Mean added weight in grams (sd) and mean percentage contribution to energy, protein and IAA from each food group across clusters.

| Cluster 1 |  |  |  |  |  |  |  |  |  |  |  |
| --- | --- | --- | --- | --- | --- | --- | --- | --- | --- | --- | --- |
| Food group | Mean weight in grams | Energy and nutrient contribution to minimum daily requirement <sup>1</sup> (%) |  |  |  |  |  |  |  |  |  |
|  | (SD) | Protein | Tryp | Thr | Leu | Lys | Met | Cys | SAA | His | Energy |
| Fruit | 90.5 (73.2) | 2.40 | 0.27 | 0.58 | 0.38 | 0.34 | 0.53 | 3.51 | 1.29 | 0.59 | 10.9 |
| Grains and pasta | 75.7 (42.5) | 21.3 | 36.1 | 23.7 | 22.8 | 13.9 | 20.8 | 55.1 | 28.6 | 28.9 | 18.8 |
| Legumes and pulses | 78.6 (48.3) | 35.3 | 30.0 | 40.5 | 31.3 | 37.9 | 21.5 | 70.5 | 33.2 | 48.1 | 11.1 |
| Nuts and seeds | 26.2 (7.92) | 24.0 | 49.3 | 33.4 | 27.4 | 18.2 | 29.6 | 58.8 | 35.4 | 40.1 | 16.2 |
| Potatoes, kumara and taro | 32.0 (29.0) | 2.75 | 6.66 | 4.25 | 2.82 | 3.86 | 3.00 | 6.07 | 3.62 | 4.09 | 9.13 |
| Sugar and sweets | 26.8 (7.98) | 5.85 | 0.81 | 0.66 | 0.61 | 0.51 | 0.58 | 1.36 | 0.75 | 0.75 | 7.41 |
| Vegetables | 26.5 (31.7) | 5.96 | 7.94 | 7.40 | 3.22 | 3.04 | 5.06 | 11.4 | 6.42 | 4.29 | 2.61 |
| Yeast and condiments | 22.1 (9.62) | 13.0 | 26.3 | 24.1 | 14.6 | 19.5 | 11.8 | 24.3 | 14.3 | 17.7 | 3.75 |
| Cluster 2 |  |  |  |  |  |  |  |  |  |  |  |
| Fruit | 86.9 (74.8) | 3.70 | 0.62 | 3.29 | 1.42 | 1.16 | 2.22 | 8.23 | 3.67 | 2.30 | 7.38 |
| Grains and pasta | 88.0 (41.9) | 19.9 | 35.6 | 24.5 | 24.0 | 12.8 | 21.5 | 56.8 | 29.5 | 29.6 | 19.0 |
| Legumes and pulses | 73.7 (54.3) | 29.7 | 23.9 | 33.1 | 25.3 | 30.9 | 17.9 | 58.9 | 27.6 | 39.2 | 9.38 |
| Nuts and seeds | 26.0 (8.23) | 21.1 | 46.5 | 29.3 | 24.9 | 16.2 | 26.3 | 51.1 | 31.2 | 35.7 | 14.0 |
| Potatoes, kumara and taro | 51.3 (26.3) | 3.53 | 8.83 | 5.76 | 3.77 | 5.00 | 3.92 | 8.08 | 4.77 | 5.40 | 10.8 |
| Sugar and sweets | 19.3 (10.9) | 2.93 | 0.80 | 0.48 | 0.48 | 0.32 | 0.46 | 1.31 | 0.65 | 0.61 | 4.95 |
| Vegetables | 54.0 (26.9) | 5.02 | 7.30 | 7.26 | 5.24 | 3.97 | 5.55 | 8.37 | 5.93 | 5.52 | 2.66 |
| Yeast and other condiments | 23.8 (10.4) | 7.26 | 12.8 | 11.4 | 6.96 | 9.33 | 5.60 | 11.3 | 6.75 | 8.36 | 1.97 |
| Cluster 3 |  |  |  |  |  |  |  |  |  |  |  |
| Fruit | 0 | 0 | 0 | 0 | 0 | 0 | 0 | 0 | 0 | 0 | 0 |
| Grains and pasta | 70.5 (40.2) | 12.8 | 21.7 | 15.7 | 14.7 | 8.76 | 13.9 | 31.7 | 17.7 | 17.7 | 16.2 |
| Legumes and pulses | 55.4 (40.9) | 25.1 | 16.1 | 26.1 | 20.0 | 25.2 | 14.0 | 50.2 | 22.7 | 32.2 | 8.05 |
| Nuts and seeds | 26.2 (7.89) | 17.7 | 33.1 | 22.4 | 20.6 | 13.0 | 18.3 | 39.1 | 22.6 | 29.7 | 13.9 |
| Potatoes, kumara and taro | 75.0 (0) | 5.36 | 14.5 | 9.23 | 6.13 | 8.38 | 6.53 | 13.2 | 7.87 | 8.9 | 15.0 |
| Sugar and sweets | 25.0 (12.2) | 3.68 | 1.37 | 0.81 | 0.82 | 0.54 | 0.77 | 2.23 | 1.11 | 1.04 | 5.91 |
| Vegetables | 3 (0) | 3.77 | 7.44 | 6.73 | 3.08 | 2.25 | 4.77 | 6.70 | 4.97 | 2.95 | 0.71 |
| Yeast and other condiments | 0 | 0 | 0 | 0 | 0 | 0 | 0 | 0 | 0 | 0 | 0 |

<sup>1</sup>Minimum daily requirement: protein was based on EAR (differentiated by sex and age), IAA was based on Table 1 and energy was based on minimum energy requirement with the consideration of PAL. Abbreviations: sd, standard deviation, Tryp, Tryptophan; Thr, Threonine; Leu, Leucine; Lys, Lysine; Met, Methionine, Cys, Cystine; SAA, sulphur amino acids; His, histidine

### 3.2 Assessment of protein and IAA intake after optimisation

Optimisation resolved all shortfalls for protein and each IAA, achieving daily requirements for all nutrients, across all three clusters for all but a few daily diets. In Figure 2, we present comparisons of the baseline and final diets for three selected IAAs, lysine, leucine and methionine. Results for energy, protein and other IAAs can be found in **Figure S1**. The violin plots illustrate the cohort distribution of nutrient intake relative to daily requirements. Requirements are demarcated at 100% of the y-axis and plot points above the 100% line indicate fulfilment of the daily requirement for the nutrient.

**Figure 2.**
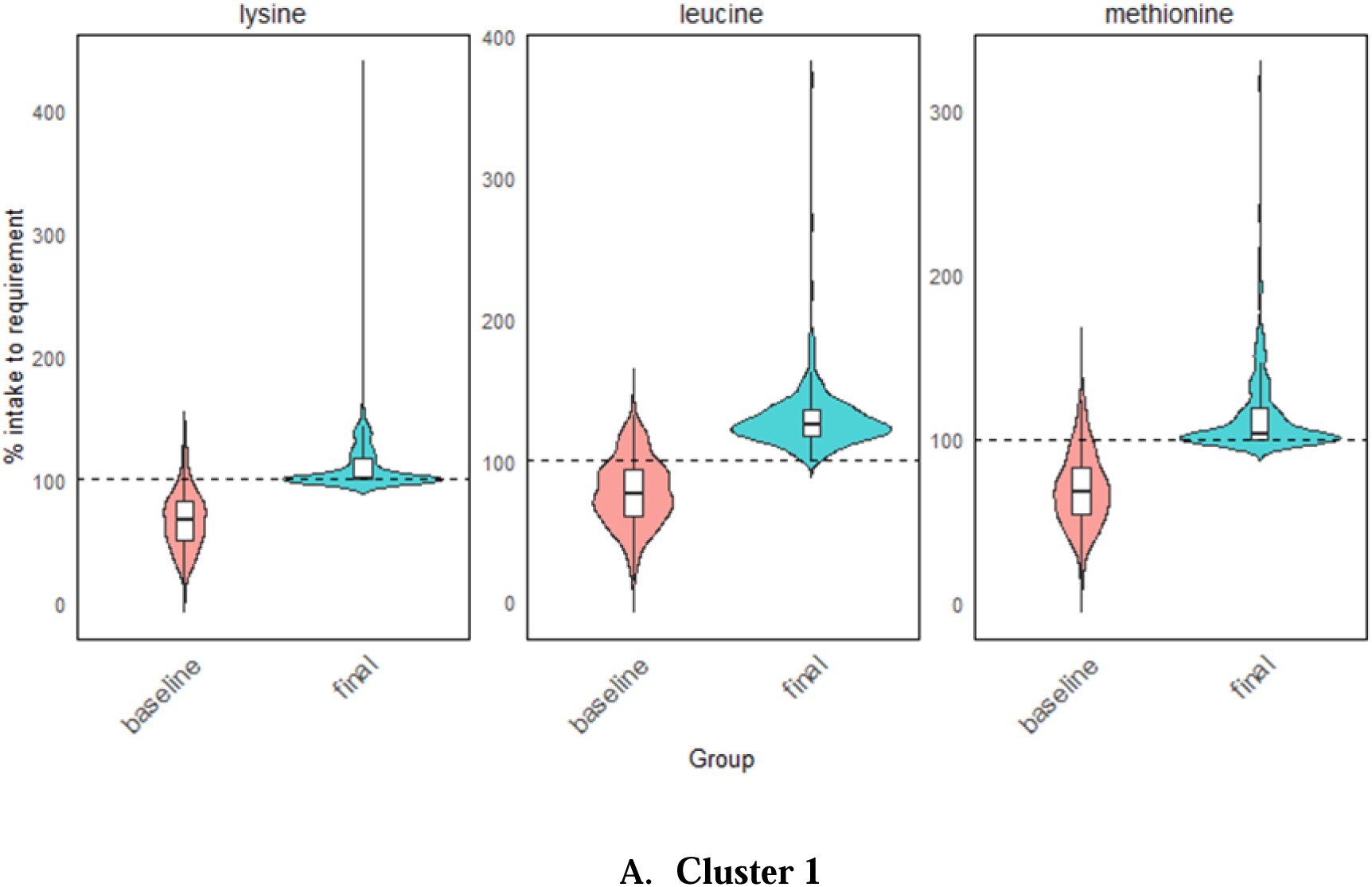
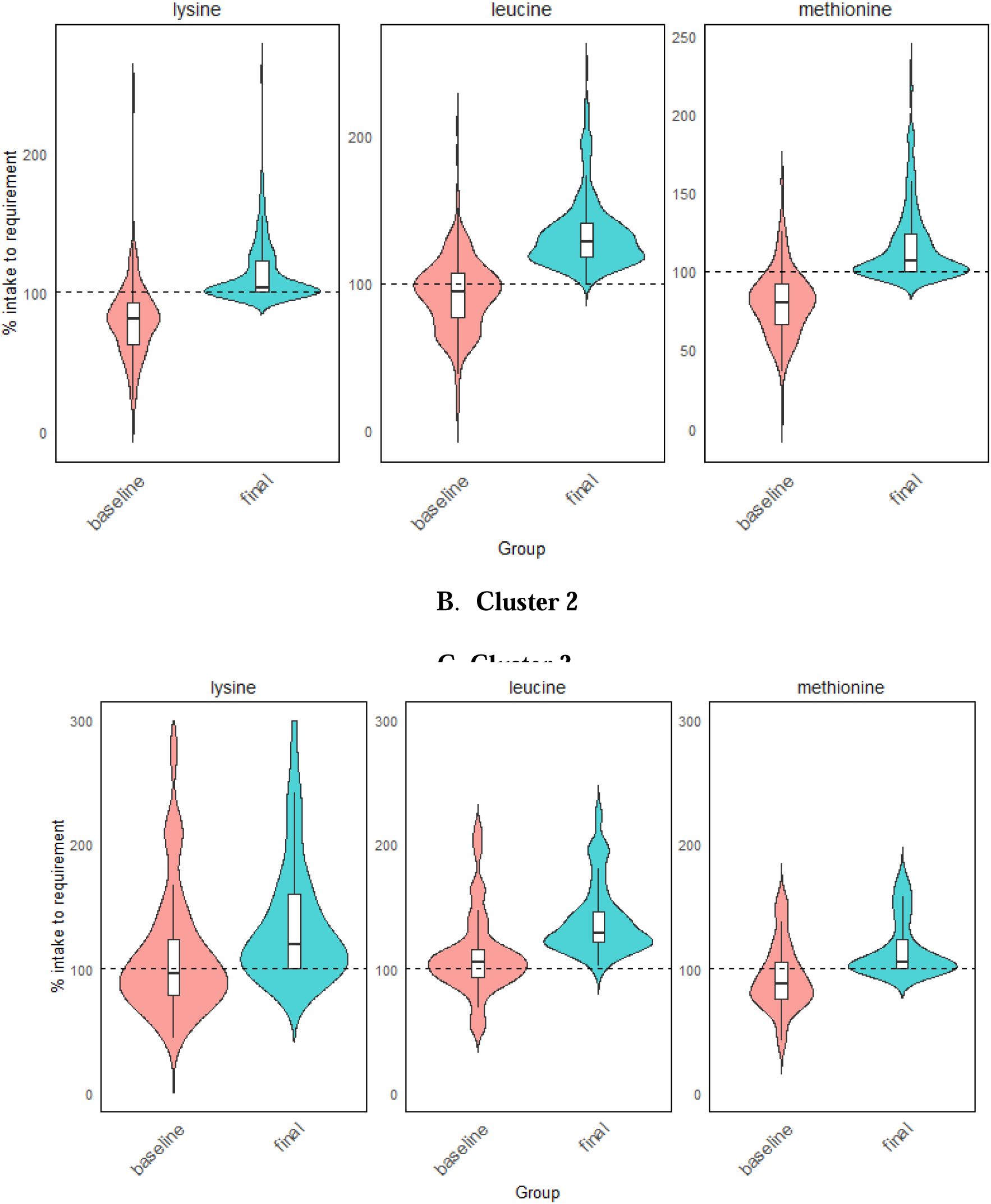
Comparison of adequacy in three selected IAAs, leucine, lysine and methionine, at baseline (unmodified diets) and final modified intakes following diet optimisation in cluster 1 (A), cluster 2 (B) and cluster 3 (C). The dotted line at 100 % indicates the average minimum daily requirement for energy and each nutrient. Each violin plot with its box plot represents the overall distribution of energy or nutrient intake in each cluster, with wider portions in the violin plot representing a larger proportion of daily diets. The top and bottom of the violin plots correspond to the maximum and minimum observed values of the cohort, smoothed by density estimate. The horizontal line in the box plot represents the median (50^th^ percentile) of the data with the interquartile ranges at 25^th^ and 75^th^ percentiles (lower and upper edges of the box plot respectively). The lines extending from the percentiles represents the range (smallest to the largest non-outlier values) which is within 1.5 times of the IQR. As the violin plots represent kernel density estimates, proportions of these plots may appear to fall below the 100% requirement. However, all successfully modified diets meet at least 100% for protein and all IAAs.

Cluster 1 had the largest proportion of baseline daily diets below requirement as compared to the other clusters. This is most apparent for nutrients like leucine (83.6%), lysine (92.7%) and methionine (90.9%). For the baseline diets in cluster 2, similarly large proportions of daily diets fell below the nutrient requirements, especially for lysine and methionine, at 84.1% and 87.1% respectively, but less so for leucine, where 62.6% were below requirements. In contrast, baseline diets in cluster 3 were closer to daily requirements for all nutrients, with fewer daily diets falling below the requirements, even for leucine (41.7%), lysine (52.8%) and methionine (69.4%).

All final diets, except for the 52 unsolved ones followed the constraints set by the LP model and achieved at least 100% of the daily required intake for protein and all IAAs. This is evident from the box plots and lower range boundaries in all three clusters which consistently met or exceeded the 100% adequacy threshold, without exceeding energy constraints (**Figure S1**). The wider base of all the final violin plots in Figure 2 indicated a higher density of daily diets with nutrient intakes that just met the minimum requirement (100%), most commonly for leucine, lysine and methionine across all the clusters, but most prominently for cluster 1, which had the highest percentage of optimised diets only just meeting the minimum requirement for these nutrients.

**Table 6** presents the mean intake of protein and IAA of all clusters relative to individual body weight at baseline and in final diets. In the final diets, the EAR was achieved for all individuals and satisfied the model constraint for protein requirement. Most individuals were within the AMDR for protein of 10 to 35%, but 14.1% and 9.2% in cluster 1 and 2 respectively were below this range, showing a smaller contribution of protein intake as a portion of daily energy. The mean daily protein consumed in grams per individual increased to 60.9 g (16.0) from 39.8 g (11.8) at baseline in cluster 1. The value was 68.6 g (16.7) in Cluster 2 compared to 51.4 g (15.3) at baseline, and 88.9 g (23.4) in Cluster 3 compared to 76.7 g (19.5) at baseline.

**Table 6.** Mean daily nutrient intake (sd) in the baseline and optimised diets, relative to individual body weight (BW) across clusters 1 to 3.

| Protein |  |  | IAAs (mg/kg BW) |  |  |  |  |  |  |  |
| --- | --- | --- | --- | --- | --- | --- | --- | --- | --- | --- |
|  | Protein<br>(g/kg BW) | AMDR in %<br>of energy<br>from protein | Cystine | Histidine | Leucine | Lysine | Methionine | SAA | Tryptophan | Threonine |
| Cluster 1 |  |  |  |  |  |  |  |  |  |  |
| Baseline | 0.59 (0.17) | 10.2 (2.43) | 6.99 (2.29) | 10.9 (3.41) | 30.2 (9.30) | 20.0 (6.99) | 6.92 (2.28) | 13.9 (4.39) | 5.03 (1.63) | 15.2 (4.75) |
| Final | 0.90 (0.23) | 12.6 (2.98) | 11.6 (3.38) | 18.4 (4.23) | 50.6 (9.75) | 33.8 (8.99) | 11.5 (2.71) | 23.1(5.73) | 8.04 (1.84) | 24.7 (4.8) |
| Cluster 2 |  |  |  |  |  |  |  |  |  |  |
| Baseline | 0.72 (0.23) | 11.7 (3.41) | 8.32 (2.53) | 13.0 (3.67) | 36.1 (10.2) | 23.8 (7.80) | 8.04 (2.21) | 16.4 (4.55) | 5.68 (1.83) | 17.5 (4.57) |
| Final | 0.96 (0.23) | 13.5 (3.45) | 11.9 (2.79) | 18.9 (3.5) | 52.2 (9.54) | 34.3 (7.03) | 11.7 (2.52) | 23.6 (4.94) | 8.07 (2.17) | 24.9 (3.8) |
| Cluster 3 |  |  |  |  |  |  |  |  |  |  |
| Baseline | 1.02 (0.31) | 17.6 (7.21) | 11.1 (3.83) | 16.8 (5.58) | 43.3 (12.9) | 33.4 (15.0) | 9.30 (2.69) | 20.3 (6.20) | 5.06 (2.07) | 21.1 (6.23) |
| Final | 1.18 (0.34) | 17.7 (4.93) | 13.5 (3.76) | 21.1 (5.17) | 54.3 (11.2) | 41.0 (14.5) | 11.6 (2.28) | 25.1(5.72) | 6.51 (1.97) | 26.2 (5.28) |

390 Abbreviations: AMDR, Acceptable Macronutrient Distribution Range, sd, standard deviation; BW, body weight; SAA, sulphur amino acids

The final IAA intake was compared to the upper limits (Table 3). Only two daily diets had an intake greater than the NOAEL for lysine but were below the LOAEL. These cases were from cluster 1 (female) and cluster 3 (male). The former case also had an elevated intake of protein relative to body weight, at 3.03 g / kg BW. This result could be attributed to a large serving (150g) of nutrient dense pea protein isolate chosen by the model, which contributed high quantities of lysine. For the latter case, protein intake was 1.67 g / kg BW. The high lysine intake could again be attributed to the addition of one more serving of pea protein during optimisation, on top of 3 servings at baseline.

The final optimised diet was compared to the baseline diet in Figure 3. A decrease in energy and nutrient contribution from fruit, vegetables and grains was observed when shifting from the baseline to final diet largely due to substantial increases in other food groups. Nuts and seeds and yeast products provided an increased contribution to energy, total protein and all IAAs from baseline to final diets (**Table S2).** In contrast, apart from increases in the contribution to protein and lysine, the nutrient contributions by legumes and pulses remain relatively unchanged in baseline and final diets (Table S2).

**Figure 3.**
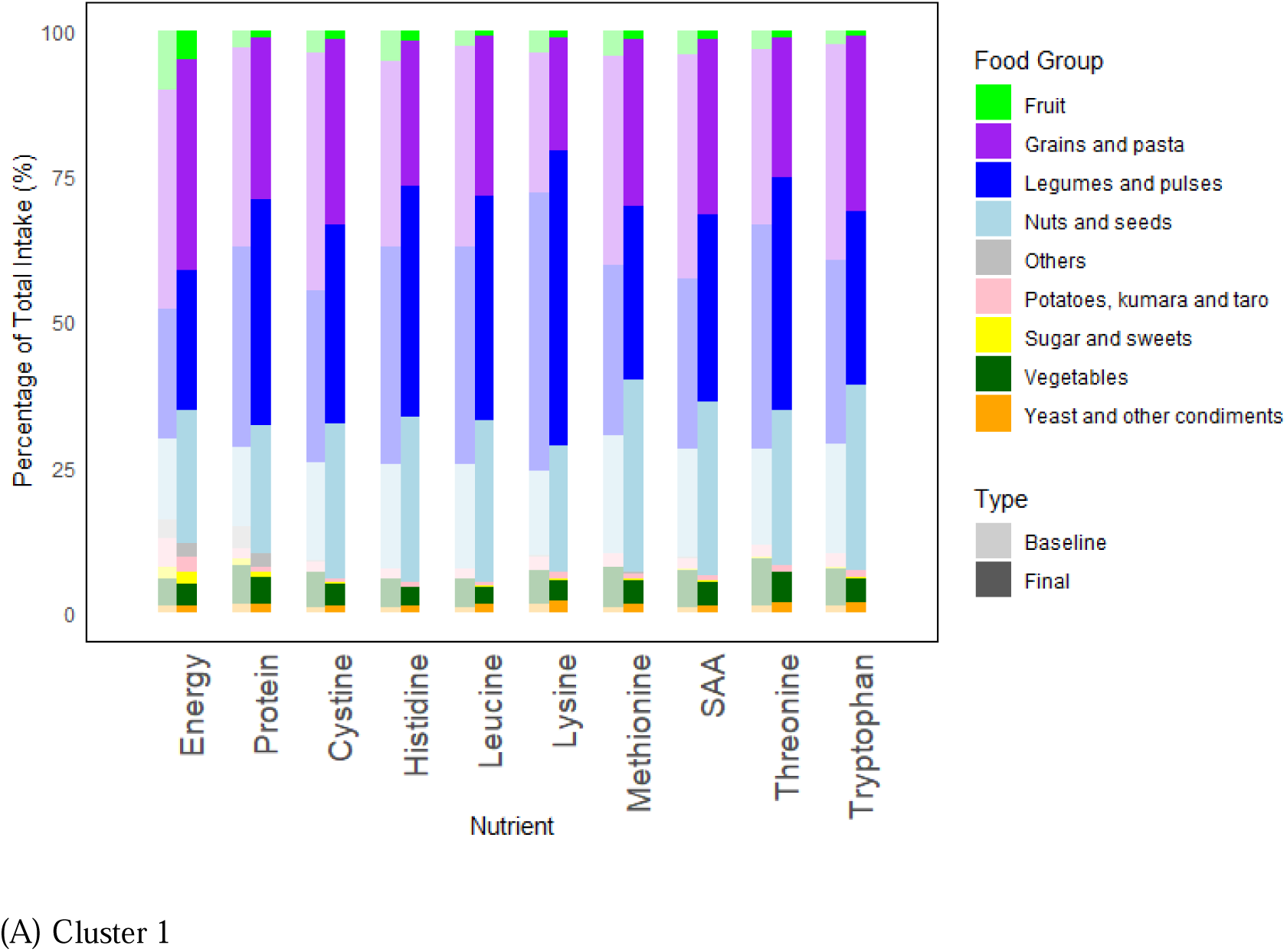
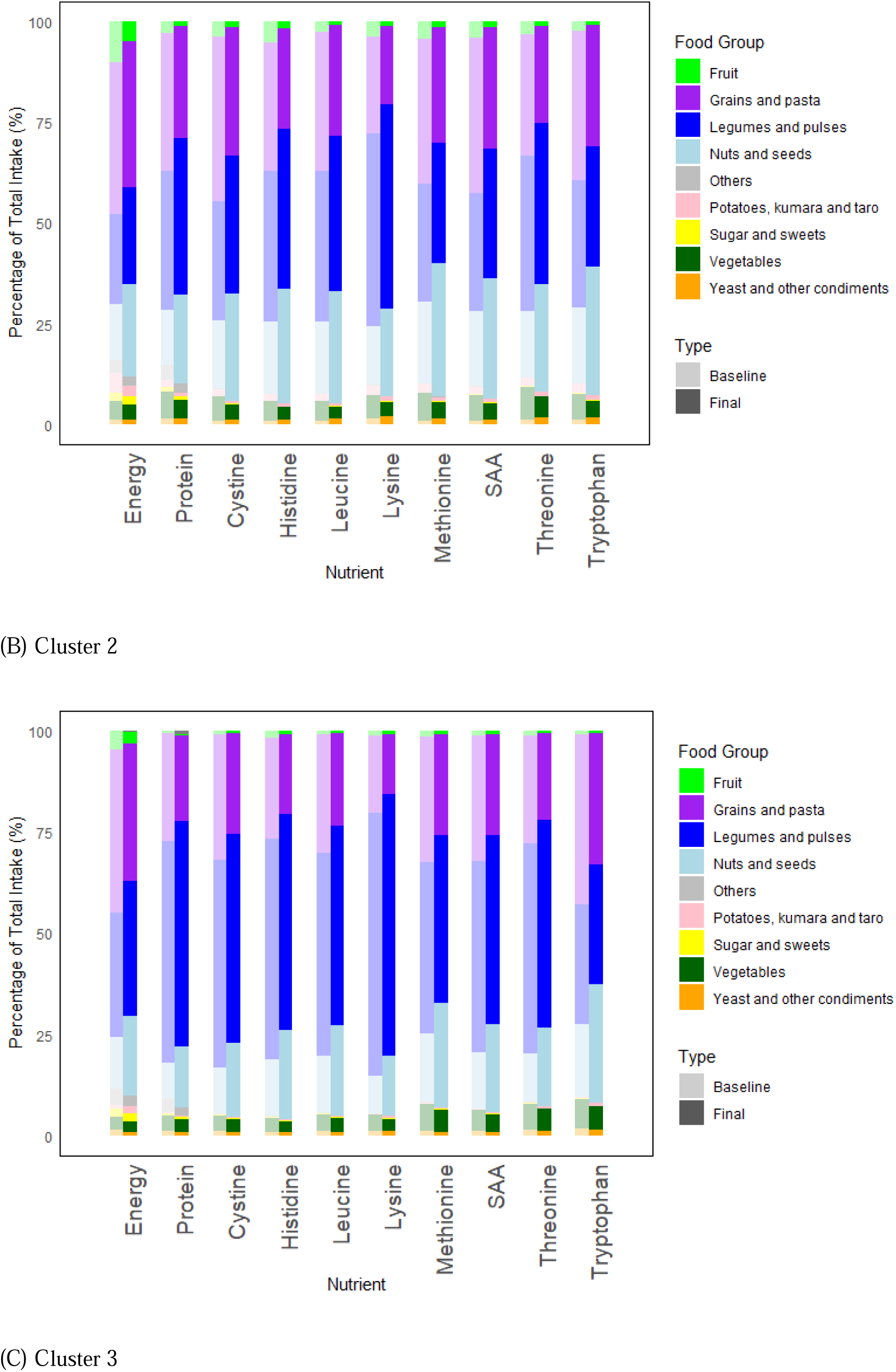
Comparison of the final modified diet with the original diet across clusters 1(A), 2(B) and 3 (C). Stacked bar graphs represent the contribution of each food group to energy, total protein and IAAs as a percentage of the total intake. “Others” consist of alcoholic and non-alcoholic beverages.

The breakdown of the proportion based on total weight of food consumed, for each food group is shown in **Table 7**. Grains and legumes contribute the largest proportion of total food mass in a roughly 1:1 ratio. When comparing how total weight of each food group changed from the baseline to final diet, food groups that had at least a 10% increase in all clusters were “legumes and pulses” and “nuts and seeds” (Table S2). Food groups that had a reduction in total weight (noting that this only occurred in cases where foods were required to be removed from the diet to provide residual energy capacity) were mainly energy-dense, from “potatoes, kumara and taro”, and “others”, which consisted of alcoholic and non-alcoholic beverages such as fruit juices and sugar-sweetened beverages (SSBs). In cluster 3, the decrease in total weight of “yeast and condiments” was due to the removal of energy-dense sauces and the increase in total weight of “potatoes, kumara and taro” occurred in two cases in cluster 3 to meet energy shortfalls.

**Table 7.** Daily mean food weight (%)^1^ contributed per food group, for each cluster.

| Food group | Cluster 1 | Cluster 2 | Cluster 3 |
| --- | --- | --- | --- |
| Fruit | 13.8 | 12.3 | 13.0 |
| Grains and pasta | 27.9 | 29.3 | 31.6 |
| Legumes and pulses | 29.3 | 31.4 | 34.1 |
| Nuts and seeds | 11.4 | 10.6 | 12.6 |
| Potatoes, kumara, taro | 11.7 | 14.1 | 5.98 |
| Vegetables | 15.2 | 15.9 | 14.4 |
| Yeast and other condiments | 3.93 | 2.50 | 3.08 |
| Sugar and sweets | 3.81 | 2.14 | 2.03 |
| Others<sup>2</sup> | 18.1 | 20.1 | 19.6 |

<sup>1</sup> Values do not sum to 100% as an aggregate of each FG across individuals of each cluster are presented.
<sup>2</sup> Others denote beverages, not including PBA that have been classified into the grains, legumes and nut (e.g. oat, almond and soy-based beverages) group

### 3.3 Sensitivity Analyses

Sensitivity analyses were conducted on the 52 daily diets which had no solutions. We assessed whether increasing the energy allowance (EER_max_) and removing serving size restrictions would improve the percentage at which each nutrient’s shortfall can be fulfilled. Even with increments in energy up to twice the EER_min_ – a level that is likely unrealistic - feasible solutions could not be achieved for most cases. Only five individual diets were able to reach at least 70% of intake for the nutrient that was in shortfall. This was only possible because these diets had fewer nutrient gaps and smaller shortfall magnitudes, making it more attainable to meet requirements within the relaxed energy constraint and limited food options available in the baseline diet.

Without serving size limits, the LP solver selects foods from the existing diets to meet nutrient targets, by adding any quantity of available dense protein sources (seeds, pulses and isolates). Low-protein foods such as vegetables were also added in larger amounts. These are unrealistic in practice to cover nutrient shortfalls. When eliminating energy and serving size constraints, most of the foods in the 52 unsolved diets are sub-optimal in reconciling protein and IAA shortfalls. Together, the exploration of these different scenarios confirm that the model formulation is robust and provided predictable changes in the outcomes when constraints were altered.

We also explored an alternative approach for the 199 problematic cases where the model could select foods from any diet in the cohort and add them to the original diets. Extruded plant proteins – notably pea, faba bean and hemp isolates – as well as spirulina and various nuts and seeds (hemp, pine nut, walnut, sesame) were frequently selected in this approach. These foods were also commonly found in the daily diets that had no protein or IAA shortfalls (n = 264). Nutrient shortfalls were resolved with small increases in mean energy intake of 0.71 (sd = 0.81) MJ with this approach.

A scenario where all isolates were removed from the model’s food selection was explored for all daily diets in this study, with the aim of understanding if that changes the number of feasible solutions and the impact on the added weight of food groups. Removal of isolates resulted in 11 more daily diets that had no feasible solutions, with 8 cases from daily diets that were meeting or above the EER_min_. This observation implies that the use of isolates was critical for these 11 daily diets to achieve protein and IAA adequacy. Among the daily diets that remained solvable, removing isolates caused small deviations in the average weight of each food group. Notable shifts in particular food-groups were likely reflections of the model’s need to substitute or reduce certain items once isolates were excluded.

When upper constraints for IAAs were introduced, no feasible solutions could be obtained for nearly all cases. The only exception was a single instance where wheat was added to contribute protein and energy, but not IAAs to a baseline diet which only had shortfalls for protein and energy. No solutions can be found for other cases as intakes of cystine and histidine are often above the upper IAA constraint even at baseline. Hence, the model could not identify suitable combinations of baseline foods that simultaneously resolve nutrient shortfalls while remaining within all imposed constraints.

### 3.4 Assessment of micronutrient intake after optimisation

Apart from protein and IAAs, several other nutrients such as vitamin B12, calcium, iron, iodine, zinc, ALA, sodium and dietary fibre were examined to evaluate how the food additions and substitutions affected their adequacy, as well as assess if the optimisation process introduced risks of exceeding the UL (**Table S1**). The comparison of the baseline and final nutrient intake following food additions are presented in **Figure S2** and the mean contribution of nutrient as a percentage of daily requirement by each added food group is presented in **Table S3.**

**Table 8** illustrates the mean intake of these nutrients and the percentage of the daily diets meeting the EAR. More than 90% of daily diets were meeting or above the daily requirements for dietary fibre, iron and zinc in all clusters. However, many optimised diets were still below the minimum intake for calcium, vitamin B12 and iodine in all clusters, suggesting that food additions based only on constraints in protein and energy content cannot correct for shortfalls in these nutrients for all diets.

**Table 8.**
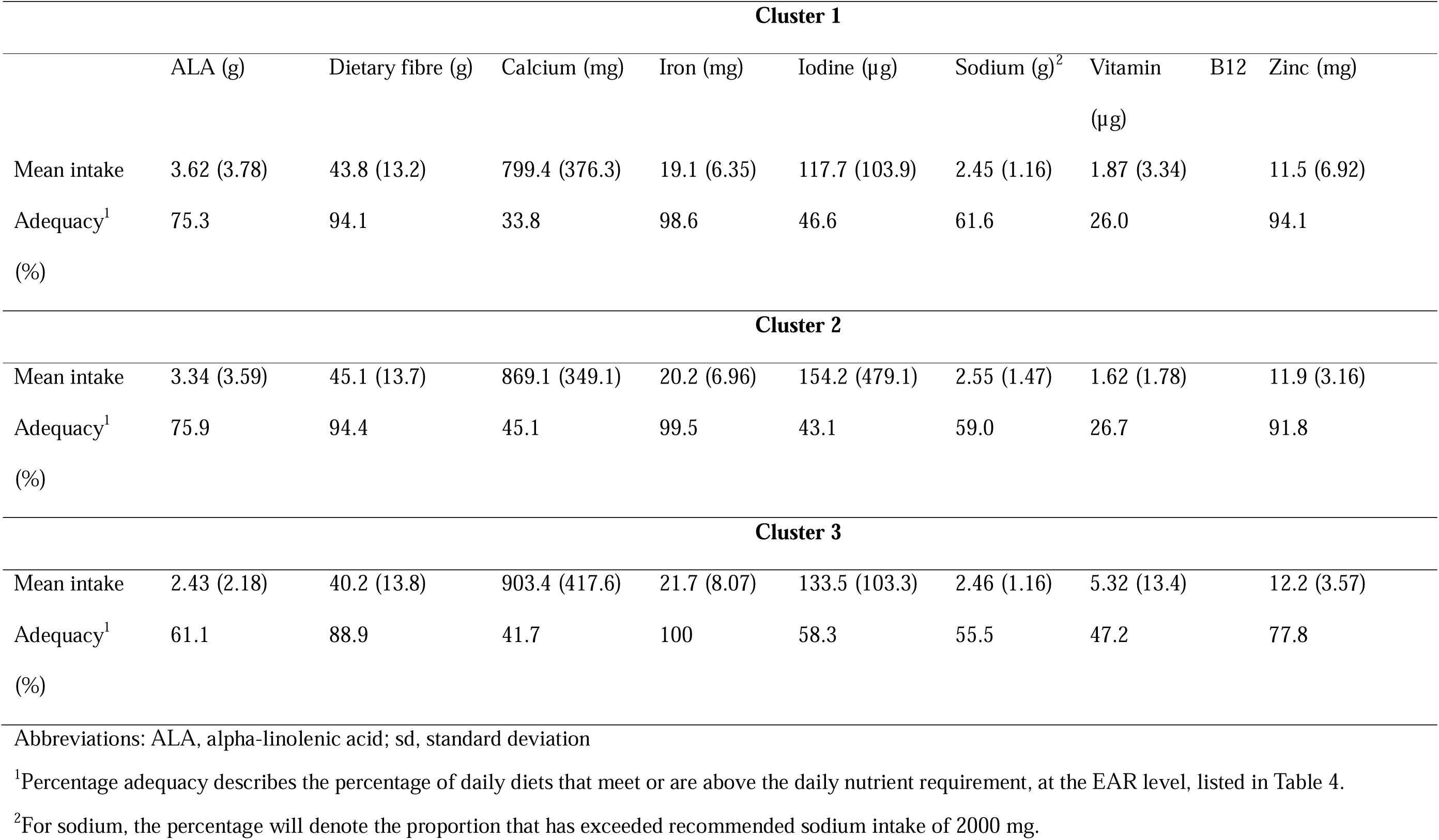
Mean intake in grams (sd) and percentage adequacy^1^ of daily diets that meet daily nutrient requirement across clusters.

A large percentage of daily diets were below the daily requirement for vitamin B12, more so for clusters 1 and 2 than cluster 3. The diets for observed outliers in each cluster with high intakes (identified in Figure S1) were examined to identify the contributing food sources. Spirulina in large quantities (15 g) present in the baseline diet, fortified yeast products, and fortified PBMA were the main food items that supplied high concentrations of vitamin B12 (**Table S3**).

The mean intakes of dietary fibre, ALA and sodium in optimised diets met the recommended daily intake (**Table 8**) across all clusters. ALA contribution was mostly from nuts and seeds, grains and pasta **(Table S3).** Sodium contributions came mainly from sauces and condiments, protein powders and PBMAs, and at least 50% of cases in the final diets, across all clusters were above the recommended intake of 2000 mg. Although other micronutrients were not prioritised in this research, **Table S4** presents the mean intake and percentage adequacy of these other micronutrients across clusters, and how these parameters change before and after diet optimisation.

Despite the mean calcium intake being close to EAR of 840 mg in all clusters, large variation existed. One case from cluster 1 was above the UL for calcium but was already above the UL at baseline. Similarly, large variations were observed for iodine with two cases from cluster 2 being above the UL but this was also observed at baseline diets of these individuals due to seaweed consumption. For zinc, one case in cluster 1 was above the UL, and was already the case at baseline due to consumption of protein powder. Three cases from cluster 2 were above the UL for iron at the final diets and were close to but not above the UL at baseline. The addition of iron-rich nuts, seeds and legumes during optimisation increased the dietary iron content.

## 4. Discussion

The main objective in this study was to address inadequate intakes in protein and IAA found in the current diets of a vegan cohort in NZ. Mathematical optimisation resolved the nutritional shortfalls of more than 90% of the daily diets while minimising the weight of food added, with the use of food items found in each individual’s existing diet. Consequently, the model generated outputs of the weight (g) and nutrient contribution for total protein, energy, IAAs, dietary fibre and micronutrients of added foods. We demonstrated the ability of the model to meet individual protein intake and protein quality requirements through the addition of food items – largely from the “legumes and pulses” and “nuts and seeds” groups. Since foods added were derived from each individual’s baseline diet, this strategy aimed for optimised diets that were more acceptable and personalised to each individual.

### 4.1 Optimising protein within energy constraints

The weight of food added must be guided by both the energy and serving size constraints as these parameters ensure that optimised diets are feasible in reality. Designing optimal vegan diets can be challenged by the need to correct nutrient shortfalls while simultaneously maintaining energy intake within limits – two constraints that may be incompatible within the baseline diet (3, 37). This was the case in this study for individuals who were already meeting or above the maximum boundary of EER but had shortfalls for protein and/or amino acids.

Substitution of foods in step 6 of Figure 1 enabled 147 daily diets, out of an unsolved 199, to meet protein and IAA shortfalls, while keeping below the EER_max_. The majority of removed foods were fruit, beverages (alcoholic, SSBs and fruit juices), grains and potatoes. Eliminating these foods provided residual energy capacity to accommodate the addition of more protein-dense foods, from legumes, pulses, nuts and seeds - food groups which had a larger increase in total weight in the optimised diets as compared to other food groups. This observation was aligned with a Swedish diet optimisation study, where pulses, meat and dairy substitutes (pea, soy and mycroprotein-based products), but not nuts, had a ten-fold increase compared to the baseline diet (12).

Legumes, nuts and seeds are noted to be good sources of lysine in a vegan diet. Since lysine was a commonly deficient amino acid in the baseline diet of this vegan cohort, the increased intake of these foods resolved this nutrient gap. Replacement of animal sources with legumes, nuts and seeds in other PB diet scenarios produced similar observations (13, 38). From a protein quality perspective, prioritising protein- and IAA-rich foods while removing energy-dense and protein-poor options such as fruits is a strategic approach to achieving the daily requirements of these nutrients within a vegan diet. However, fruits and vegetables are an excellent source of vitamin C (39) and other essential micronutrients, and are central components of Food Based Dietary Guidelines (FBDGs) around the world (40, 41). Therefore, while optimising for protein adequacy, dietary modifications must still consider the roles of these foods in ensuring overall nutritional balance.

While nuts and seeds are energy-dense foods, they were favoured by the LP algorithm and increased in the final diets, most prominently for clusters 1 and 2. When comparing the current Danish diet with the *NutriHealthGHGE* diet - an optimised diet modelled to meet nutritional adequacy, health and greenhouse gas emissions (GHGE), substantial shifts in protein sources were observed - ruminant meat was removed, other animal-sourced proteins were reduced and nut intake increased by 230% (42). Nuts and seeds were observed as a key source of total protein, leucine and methionine in our optimised diets, contributing higher quantities of these nutrients per gram of food when compared to legumes and pulses. Nuts as valuable protein sources were highlighted in food replacement scenarios among pregnant women in Australia and identified to contribute at least twice the protein of tofu, legumes and pulses (43). Although the study did not account for protein digestibility, nuts was the only PB food group identified in the study to contain higher protein content than an isoenergetic serve of an animal-sourced food. Nuts and seeds were commonly consumed in the baseline diet of this vegan cohort, another reason that could drive their selection in the optimised diets.

On average in the optimised diets, the mean intake per individual across the whole vegan cohort for legumes and pulses was 129g, for nuts and seeds was 38 g and for grains was 108g. This was roughly equivalent to the quantity of protein-rich foods (consisting of beans, peas, lentils, soy products, nuts and seeds) in a modelled 2000 kcal-per-day vegan diet in America, but the quantity of grain was higher at 184 g per day (44). This difference could be attributed to the narrower individual energy boundaries in our study cohort, leading to the model favouring less energy-dense but protein rich foods. The modelled American vegan diet met the reference value for protein, but amino acid intake was not analysed, so the proportion of these foods may require adjustments if protein quality is included. For example, another diet simulation on American adults found that the IAA scores increased when 50% of amino acids from grains were replaced by amino acids from lentils and legumes (45).

Given the high protein density and protein quality of legumes, nuts and seeds, and their complementary effect with grains to achieve protein adequacy, the proportion and balance of these foods within the contexts of vegan diets warrant further investigation. Subsequent updating of serving size guidelines in FBDGs such as the Eating and Activity Guidelines for NZ adults (28) could provide clearer guidance for vegan consumers in meeting all IAA requirements. The best ratio of grain to legume in plant-based protein complementation remains unclear and will vary across different foods. For example, substituting 200g of cooked chickpeas with 120 g of cooked lentils improves the protein quality score due to higher protein content of lentils (46) showing variation in protein quality even within a food group. Our study found an approximate 1:1 ratio but a 2:1 ratio may be more effective for optimal protein quality (47). More experimental verification or modelling at the meal level is necessary to establish more precise ratios of these foods.

Extrusion processes in the manufacture of protein isolates improve protein digestibility by reducing the anti-nutritional factors in these foods (48, 49). Hence, their addition to a vegan diet could be beneficial in promoting protein adequacy. However, as compared to nuts and seeds, which are versatile additions to meals and snacks, protein isolates and spirulina are more challenging to incorporate into vegan diets without adding sweeteners - such as fruit or PB beverages - to enhance palatability. We highlight this as an opportunity for innovation in the formulation of PB meat and dairy alternatives, where the integration of high-quality protein isolates as ingredients can improve nutrient shortfalls in an acceptable form. The value of these isolates was evident in the sensitivity analyses of the 52 unsolved cases, where their addition to the baseline diet was more effective in fulfilling shortfalls than restricting the model to add or substitute foods that was present in the individual’s diet.

### 4.2 Evaluation of optimised diets for dietary fibre and micronutrients

The post-optimisation analysis of dietary fibre, ALA, calcium, zinc, iron, iodine, sodium and vitamin B12 were examined. The most concerning nutrients at risks of deficiencies were vitamin B12, iodine and calcium, where the percentage of daily diets achieving adequacy was approximately 50% or below. Results were generally poorer in clusters 1 and 2.

Small quantities of vitamin B12 can be found in the baseline diets of foods consumed by this vegan cohort, such as seaweed, protein powder and yeast products (50). Spirulina - a food selected in some instances by our model - is a rich source of vitamin B12 (29) and contained the largest quantity of the micronutrient at 3μg per gram in our vegan dietary records. It is also a high contributor of iodine (22μg per gram). However, spirulina is not commonly selected due to its infrequent consumption in the baseline diet. As bread is mandatorily fortified with iodine in NZ (51), the addition of bread, mostly of the multigrain and seeded varieties, contributed varied quantities of iodine, alongside protein and IAAs in the optimised diets. Calcium-rich foods in our dietary data were tofu, soy-based PB food and beverages, chia and sesame seeds. These foods are not only effective protein sources that contribute substantially to digestible total protein and IAAs but may have value in increasing calcium intake in a vegan diet. The higher calcium content of PB alternatives may be related to calcium fortification of these foods. However, lower and more variated calcium content exists across these alternatives as compared to animal-sourced foods like cow’s milk (52, 53). Furthermore, the quantity of utilisable calcium from PB foods will be reduced due to the presence of anti-nutritional factors that impair bioavailability (54). The inclusion of soy products and some types of seeds can improve the protein quality and calcium content of the vegan diet but may be infrequently selected by our model if these foods are uncommon or absent in existing diets.

The primary constraint of optimisation was to fulfil total protein and protein quality targets with minimal food volume added. Hence, the model will not allocate additional quantity of food to address micronutrient shortfalls as they fall outside the optimisation targets. Furthermore, fortified foods such as PB beverages were not prioritised in the selection process due to their poorer protein quality and higher energy content (55). However, fortified foods have an important role in micronutrient contribution in a vegan diet. For example, the selection of a fortified soy drink in a vegan diet scenario in The Netherlands increased calcium intake to requirements (40). Similarly, nutritionally adequate optimised diets in a vegan Swedish diet that met calcium and vitamin B12 requirements was plausible due to the inclusion of high quantities of fortified PB alternatives (12).

If micronutrient constraints are to be included in our optimisation process, the number of feasible solutions would likely decrease as fewer foods within existing PB diets would meet protein quality and all micronutrient requirements within the required energy boundary. Deficits in these micronutrients could also be related to the exclusion of supplements in the modelling process and thus an underestimation of total micronutrient intake is reflected. Nevertheless, our analysis suggests that despite the ability to achieve protein intake and protein quality requirements in the vegan diet of the NZ cohort while keeping to energy constraints, ensuring the intake of all micronutrients, especially for calcium, vitamin B12 and iodine from food sources alone remains a challenge. Solutions that can minimise energy addition may require replacing foods with fortified versions or adding supplementation to the diet. In support of this, Storz et al (2023) observed adequate vitamin B12 status among vegans due largely to > 90% of cohort consuming a daily median intake of 250 μg vitamin B12 supplement (56).

Elevated nutrient intake due to the addition of legumes, nuts and seeds in the optimised diets contributed strongly to ALA, iron and zinc (43, 57), and in three cases in clusters 2, propelled iron intake to above the upper boundary of the individuals. However, we have not accounted for the bioavailability of iron and zinc in the modelled diets and the requirements for these nutrients may be elevated in a vegan diet to accommodate poorer bioavailability.

Across all clusters, a high intake of dietary fibre was also introduced in the optimised diets, with more than 90% of individuals in all clusters consuming quantities above the requirements. While dietary fibre intake is associated with several health benefits including enhanced satiety which has a beneficial role in appetite regulation, it can impair nutrient digestion and utilisation by the body (58). The introduction of large quantities of dietary fibre in our modelled diets may present a practical challenge in reality as overall food intake may become limited due to satiety (43, 59). Hence, this may restrict the consumption of protein, IAAs and essential micronutrients.

Another conundrum in vegan dietary planning is related to the introduction of novel PBAs which are potentially good sources of digestible protein but could inevitably raise sodium intake (60, 61). In our modelled diets, the addition of PBAs, snack foods and soy-based condiments raised the sodium intake. However, at baseline, approximately 50% of individuals in each cluster were already consuming sodium above the requirements. Several health concerns are associated with extended periods of high sodium intake such as hypertension and cardiovascular diseases (62). The selection of traditional protein sources such as beans and lentils in optimised diets would likely contribute less to sodium intake than the more processed alternatives (61).

### 4.3 Strengths and limitations

A key strength of this study lies in the use of individual-level optimisation, incorporating personalised calculations of energy, total protein and IAA requirements based on each participant’s body weight, sex, age and physical activity. This enabled the identification of specific nutrient shortfalls for targeted optimisation. Compared to population-level modelling, this individualised approach allows for more personalised nutritional strategies, potentially improving the acceptability and adherence to dietary interventions aimed at improving the protein quality of vegan diets (63). Nonetheless, the limitation of the model is its dependence on food availability within the individual’s existing dietary records. This contrasts with benchmarking approaches that optimise diets by drawing from peer diets that demonstrate more favourable balance of “more-is-better” versus “less-is-better” nutrients (4). By leveraging the dietary habits of peers within the same population, such approaches enable modifications of less optimal diets using culturally appropriate alternatives to improve both nutritional quality and feasibility (4, 9). Applying this strategy in the context of vegan diets may be challenging due to limited availability of diverse, nutritionally adequate vegan dietary records in the population to serve as peer benchmarks.

A second strength in our modelling approach is the consideration of diet variety by introducing maximum serving limits of each food group, as guided by NZ dietary guidelines (28) and disallowing duplicated selections for each food item. A similar approach that applied serving size constraints, as guided by the Australian Dietary Guidelines (22), was adopted in an LP optimisation exercise for various diet scenarios in Australia (6). By placing limits on the daily intake of each food item, the model ensured that some diversity was introduced to the diet while keeping to a practical volume of food item. The model chooses up to 150 g for legumes and pulses (Table 2), but this serving size constraint may not be applicable for legume-based protein powders which are typically consumed in quantities of 20 to 35 g per serve (64). This suggests that more specific serving size guidelines for these foods is required for optimisation modelling. Another strength of this study is the adjustment for protein and IAA digestibility of plant foods, with currently available TID coefficients, thus providing a more accurate estimation of the quantity of these utilisable nutrients in the diet.

One limitation observed in the current model is that during substitutions, entire baseline food items are removed rather than having their portion sizes reduced. This means that all nutrients contributed by that food are also eliminated. Although the model may introduce the same food at a smaller serving size, perhaps a more effective approach might involve modifying the food’s recipe or formulation to improve the protein quality, rather than removing it completely. Among the 199 problematic cases, 41 cases which had feasible solutions ended up with diets that were below the EER_min_ (**Figure S2, energy panel)**. This suggests that removing the top 25% and 50% of energy-dense foods may be too restrictive in these scenarios, so enabling the partial removal rather than full elimination of some foods could be a more intuitive and realistic approach. However, introducing a solver to intuitively reduce the portions of these foods may involve longer run times and could generate unrealistic proportions of foods that do not reflect realistic quantities consumed by individuals, unless further serving constraints were implemented. While the 25% and 50% thresholds used were arbitrary, further empirical testing is needed to determine optimal cut-off points for dietary feasibility.

The feasibility of our models follows the assumption that the added foods will be acceptable because they are obtained from the current diets of each individual (65). Additionally, the solutions are only reliable when dietary intake data is reported accurately but misreporting is a common limitation with the use of dietary records. There is no measurement to determine satiety in our model from the consumption of protein and fibre-dense foods (59, 66, 67), or validate the extent of acceptability by the cohort. Furthermore, the calculation of the optimal nutrient intakes by LP is dependent on the quality of the food and nutrient composition databases used in the optimisation (6, 37). Food items from the four-day food diaries may not be an exact match to the USDA food items used, and even with normalisation will result in amino acid compositions that are not precise. In the absence of NZ amino acid composition data, this appears the best substitute. We acknowledge that defining “currently consumed foods” from the four-day records may underestimate the variety of foods consumed by individuals over longer periods. Yet, foods outside of the records does not guarantee acceptability. The vegan diet is inherently restrictive in high quality protein sources and using the dataset available captured all major food groups that feature in these diets. An alternative approach was explored where the model could draw from all available food sources. However, the optimisation consistently defaulted to a narrow set of food items for inclusion into everyone’s existing diets, such as isolates to maximise protein quality with small increases to energy, but this offers an oversimplistic solution and lack of diversity.

This study has optimised protein intake and protein quality with respect to daily requirement but has not considered how foods can be spread across the day. Distributing protein-dense foods in appropriate quantities across multiple eating occasions is better tolerated than consuming large quantities of these foods in one meal. Our future work will consider the planning of vegan meals across eating occasions of the day. This is a relevant area of study given the time-sensitive nature of amino acid metabolism, where the coordinated intake of diverse protein sources within defined temporal windows is crucial to allow synchronised availability of all amino acids, for maximum metabolic utilisation (68, 69). To do so, appropriate meal reference guidelines for protein and IAA requirements are necessary, but this information is currently lacking in the literature. Nevertheless, it is important to establish the proportion of complementary plant foods that should be included per meal and calculate the quantities of IAAs that together, meet daily intake requirements.

## 5. Conclusion

This study demonstrated the feasibility of improving the protein adequacy of vegan diets using addition and substitution strategies with mathematical optimisation. Constraining the model to select foods that already exist within each individual’s diet goes some way to ensuring acceptability. Nuts, seeds, legumes, pulses and yeast products emerged as superior sources of total protein, leucine, methionine and lysine - the most commonly limiting IAAs in the vegan diet of this cohort. The optimisation outcomes were dependent on the composition of each individual’s initial diet thus confirming our hypothesis. When large nutrient deficits were present, the model had limited ability to resolve them. We observe this as a key challenge in designing protein adequate vegan dietary patterns and the presence or absence of high-quality protein (such as isolated plant proteins) can influence the success of dietary optimisation.

Eliminating all animal-sourced foods requires careful selection of diverse plant sources to meet protein adequacy. The consequence of prolonged deficiencies in the essential amino acids can manifest in various metabolic disorders. While this study has shown how to improve protein adequacy for most individuals in the cohort, the shortfalls for calcium, vitamin B12 and iodine were not resolved from food additions and substitutions, showing that apart from protein quality, several nutrients are problematic in vegan diets and should not be neglected in diet planning. Food fortification and supplementation have key roles in reducing the risks of micronutrient deficiencies and may be more relevant for vulnerable populations like the elderly and women of reproductive age who have increased nutrient needs.

To ensure nutritional balance in vegan diets, different PB proteins need to be combined. Our modelling outcomes show that improvement in protein quality can be achieved with higher proportions of nuts, seeds, legumes and pulses to grains and other energy-dense PB foods. These conclusions could be further applied in the formulation of PB alternatives and inform dietary guidelines for vegans but require extensive verification in the actual population. Given the effect of time and distribution of protein intake on metabolic function, adequate balance is necessary across eating occasions of the day. Thus, further extensions in our modelling work are needed to derive more precise proportions of different plant-based foods within a meal context.

## Abbreviations

AA: Amino acid
AI: Adequate intake
ALA: alpha-linolenic acid
AMDR: Acceptable Macronutrient Distribution Range
BW: Body weight
Cys: cystine
EAR: Estimated average requirement
EER: Estimated Energy Requirement
FAO: Food and Agricultural Organisation
FBDG: Food based dietary guidelines
GHGE: Greenhouse gas emissions
His: histidine
IAA: Indispensable amino acid
IPAQ: International Physical Activity Questionnaire
IQR: Interquartile range
Leu: leucine
LP: Linear programming
LOAEL: Lowest-observed-adverse-effect
Lys: lysine
Met: methionine
MET: metabolic equivalent task
MJ: megajoules
NRV: Nutrient reference values
NZ: New Zealand
NOAEL: No-observed-adverse-effect-level
PAL: physical activity level
PB: Plant-based
PBA: Plant-based alternative
PBMA: Plant-based meat alternative
SAA: sulphur amino acids
SD: standard deviation
TID: True ileal digestibility
Thr: threonine
Tryp: tryptophan
UL: upper level
USDA: United States Department of Agriculture

## Data Availability

All data produced in the present work are contained in the manuscript

Dietary data of individual vegans will not be made available due to privacy concerns as food records may serve as identification of participants. Codes for the optimisation will however be made available upon request.

## Acknowledgements

The authors would like to acknowledge the Vegan Health Research Project team in Massey University, Albany for their role in managing and conducting the cross-sectional study which provided the dietary and anthropometric data for this research. We thank Professor Kathryn Beck, Professor Cathryn Conlon, Dr Hajar Mazahery, Ms Rebecca Paul, Mr Owen Mugridge. We also thank Dr Karen Mumme and the Master of Dietetics students for their role in matching and compiling nutrient content of all food items from the food diaries to the NZ food composition database. We would also like to thank Mr Jacob Knight, a Sustainable Nutrition Initiative ™ summer internship student, for his contribution to matching food items to the USDA food composition database. BXPS, MV, NS designed the research; BXPS conducted the research; BXPS, MV, NS analysed the data; BXPS wrote the paper and had primary responsibility for the final content. MV, NS, PvH, WM reviewed and edited the paper. All authors read and approved the final manuscript.

## Declaration of Generative AI and AI-assisted technologies in the writing process

Large language model (Open AI) was used to support the production of the codes used in optimisation.

