## Supplementary Information for "Moving from shortfall towards adequacy: improving the protein quality of New Zealand vegan diets through optimisation modelling"

**Table S1**. Minimum daily requirements and upper limits of nutrients

| Nutrient | Sex | Age | AI / EAR per day | UL / day |
| --- | --- | --- | --- | --- |
| ALA | Male | 19 and above | 1.3 g | - |
|  | Female | 19 and above | 0.8 | - |
| Dietary fibre | Male | 19 and above  19 and above | 30 g | - |
|  | Female |  | 25g | - |
| Vitamin B12 | Male and female | 19 and above | 2.0 µg | - |
| Calcium | Male | 19 to 70 | 840 mg | 2500 mg |
|  |  | Above 70 | 1100 mg |  |
|  | Female | 19 to 50 | 840 mg |  |
|  |  | Above 51 | 1100 mg |  |
| Iodine | Male and female | 19 and above | 100 µg | 1100 µg |
| Iron | Male | 19 and above | 6 mg | 45 mg |
|  | Female | 19 to 50  Above 51 | 8 mg |  |
|  |  |  | 5 mg |  |
| Sodium | Male and female | 18 and above | 2000 mg | - |
| Zinc | Male | 19 and above | 12 mg | 40 mg |
|  | Female |  | 6.5 mg |  |

Abbreviations: AI, adequate intake; ALA, alpha-linolenic acid; EAR, estimated average requirement; UL, tolerable upper intake level

- indicates no UL can be set for that nutrient

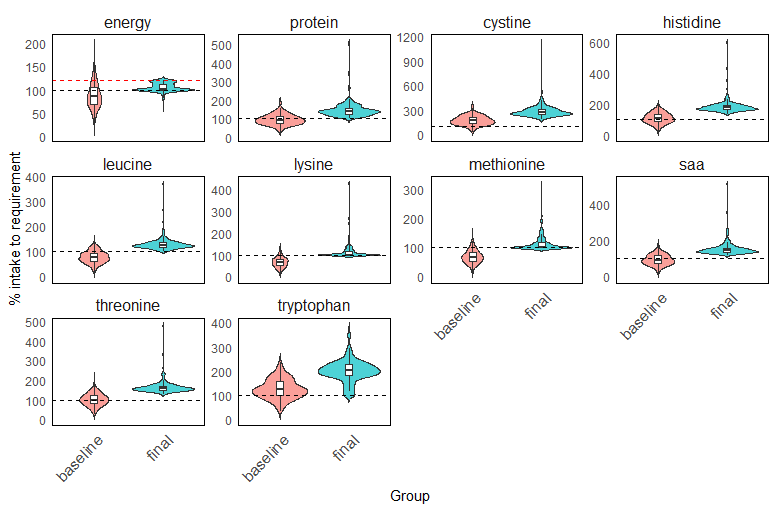

A. **Cluster 1**

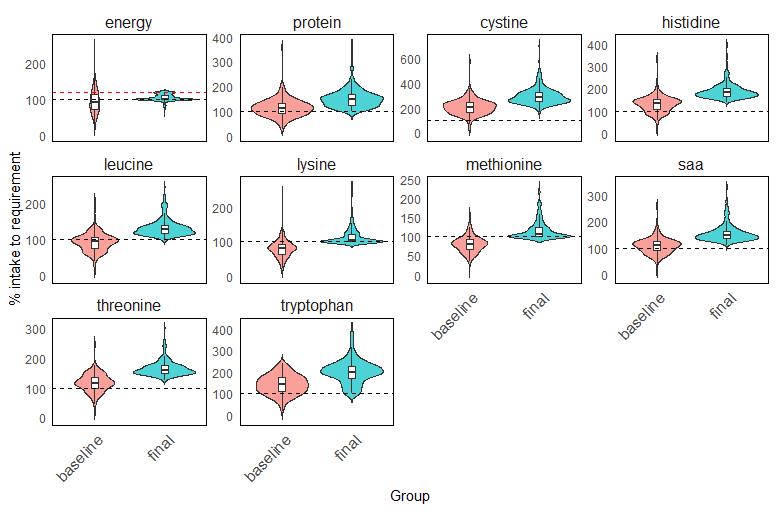

B. **Cluster 2**

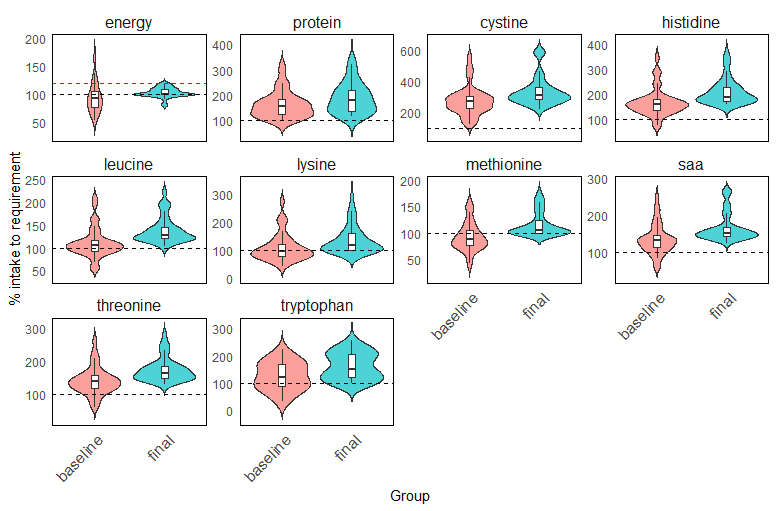

C. **Cluster 3**

**Figure S1.** Comparison of adequacy in energy, total protein and each IAA at baseline (unmodified diets) and final modified intakes following diet optimisation in cluster 1 (A), cluster 2 (B) and cluster 3(C). The dotted line at 100 % indicates the average minimum daily requirement for energy and each nutrient. The dotted red line at 120% in the energy plot indicates the average maximum daily requirement for energy. Each violin plot with its box plot represents the overall distribution of energy or nutrient intake in each cluster, with wider portions in the violin plot representing larger proportion of daily diets. The top and bottom of the violin plots correspond to the maximum and minimum observed values of the cohort, smoothed by density estimate. The horizontal line in the box plot represents the median (50^th^ percentile) of the data with the interquartile ranges at 25^th^ and 75^th^ percentiles (lower and upper edges of the box plot respectively). The lines extending from the percentiles represents the range (smallest to the largest non-outlier values) which is within 1.5 times of the IQR.

**Table S3.** Mean contribution of dietary fibre, ALA and micronutrients (%) for each added food group added after optimisation

| Cluster 1 | | | | | | | | |
| --- | --- | --- | --- | --- | --- | --- | --- | --- |
| Nutrient contribution to daily requirement ^1^ (%) | | | | | | | | |
| Food group | ALA | Dietary fibre | Calcium | Iron | Iodine | Sodium | Vitamin B12 | Zinc |
| Fruit | 0 | 19.3 | 8.46 | 33.6 | 0.29 | 1.85 | 0 | 4.76 |
| Grains and pasta | 30.5 | 18.7 | 6.73 | 29.3 | 18.3 | 13.6 | 2.84 | 19.2 |
| Legumes and pulses | 41.2 | 14.3 | 10.8 | 38.2 | 7.96 | 16.2 | 14.9 | 23.7 |
| Nuts and seeds | 144.0 | 12.3 | 4.14 | 21.2 | 0.98 | 1.28 | 0.91 | 15.1 |
| Potatoes, kumara and taro | 3.69 | 4.16 | 0.75 | 5.43 | 1.68 | 13.0 | 0 | 3.72 |
| Sugar and sweets | 1.99 | 4.52 | 1.60 | 19.4 | 1.24 | 0.35 | 1.54 | 7.44 |
| Vegetables | 5.27 | 9.31 | 2.59 | 18.6 | 28.1 | 1.41 | 196.3 | 27.6 |
| Yeast and condiments | 0.17 | 11.0 | 1.63 | 17.7 | 2.42 | 14.1 | 0 | 26.1 |
| Cluster 2 | | | | | | | | |
| Fruit | 7.33 | 18.2 | 7.69 | 25.2 | 0.04 | 3.85 | 0 | 2.32 |
| Grains and pasta | 15.7 | 15.8 | 5.99 | 28.0 | 12.5 | 13.1 | 1.10 | 14.3 |
| Legumes and pulses | 29.3 | 12.9 | 11.6 | 41.3 | 9.53 | 14.4 | 15.6 | 18.9 |
| Nuts and seeds | 102.1 | 10.3 | 3.73 | 21.6 | 1.22 | 1.40 | 0.76 | 14.8 |
| Potatoes, kumara and taro | 4.55 | 6.08 | 1.19 | 8.66 | 1.95 | 10.9 | 0 | 4.83 |
| Sugar and sweets | 1.56 | 4.08 | 1.29 | 21.6 | 1.03 | 0.19 | 1.43 | 5.50 |
| Vegetables | 13.5 | 5.61 | 3.03 | 9.00 | 4.68 | 3.16 | 26.2 | 4.17 |
| Yeast and condiments | 0 | 7.05 | 1.04 | 46.6 | 1.25 | 22.3 | 33.6 | 13.6 |
| Cluster 3 | | | | | | | | |
| Grains and pasta | 24.2 | 15.2 | 4.21 | 17.3 | 6.89 | 8.90 | 2.80 | 12.6 |
| Legumes and pulses | 22.8 | 9.11 | 10.2 | 34.3 | 7.97 | 12.4 | 9.20 | 12.0 |
| Nuts and seeds | 65.4 | 9.73 | 3.64 | 15.3 | 0.98 | 1.40 | 0 | 12.6 |
| Potatoes, kumara and taro | 5.77 | 8.50 | 1.92 | 13.8 | 3.90 | 26.4 | 0 | 5.31 |
| Sugar and sweets | 3.75 | 7.73 | 1.84 | 34.0 | 1.84 | 0.42 | 2.25 | 7.60 |
| Vegetables | 0 | 1.50 | 0.55 | 17.8 | 66.0 | 1.58 | 471.0 | 0.92 |

Nutrient contribution (%) is calculated as the total intake of each nutrient per food group as a percentage of the daily nutrient requirement (Table 4) at the EAR level.

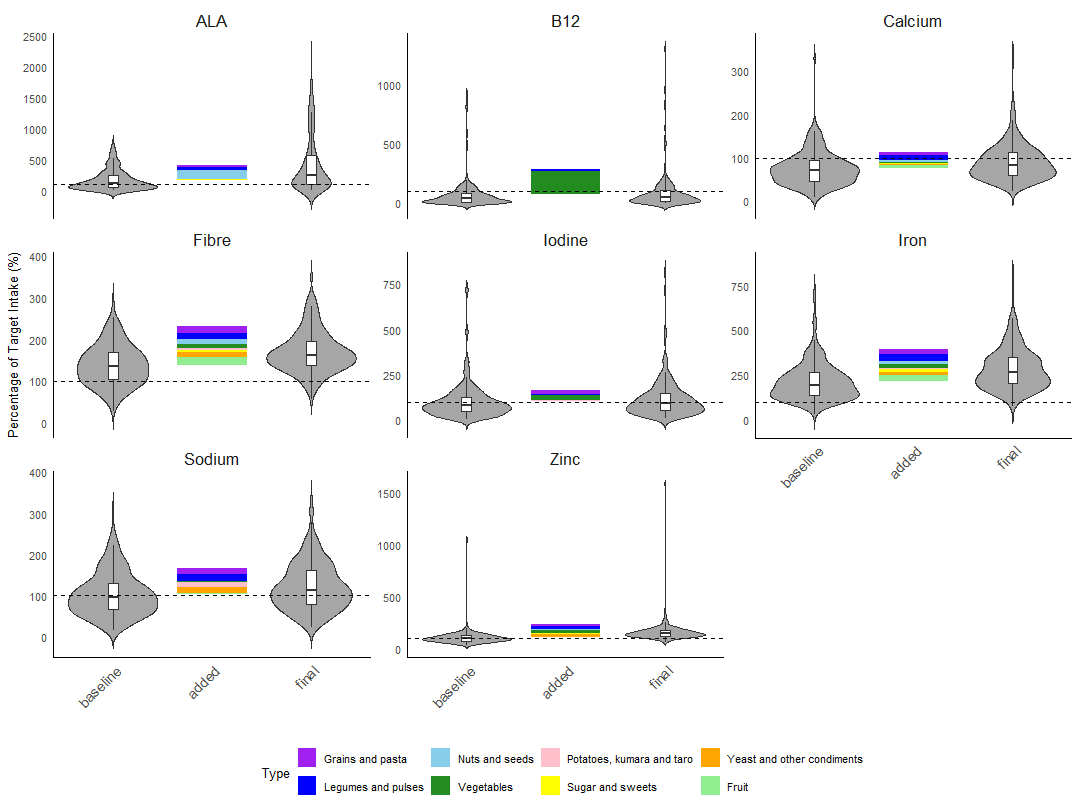

D. **Cluster 1**

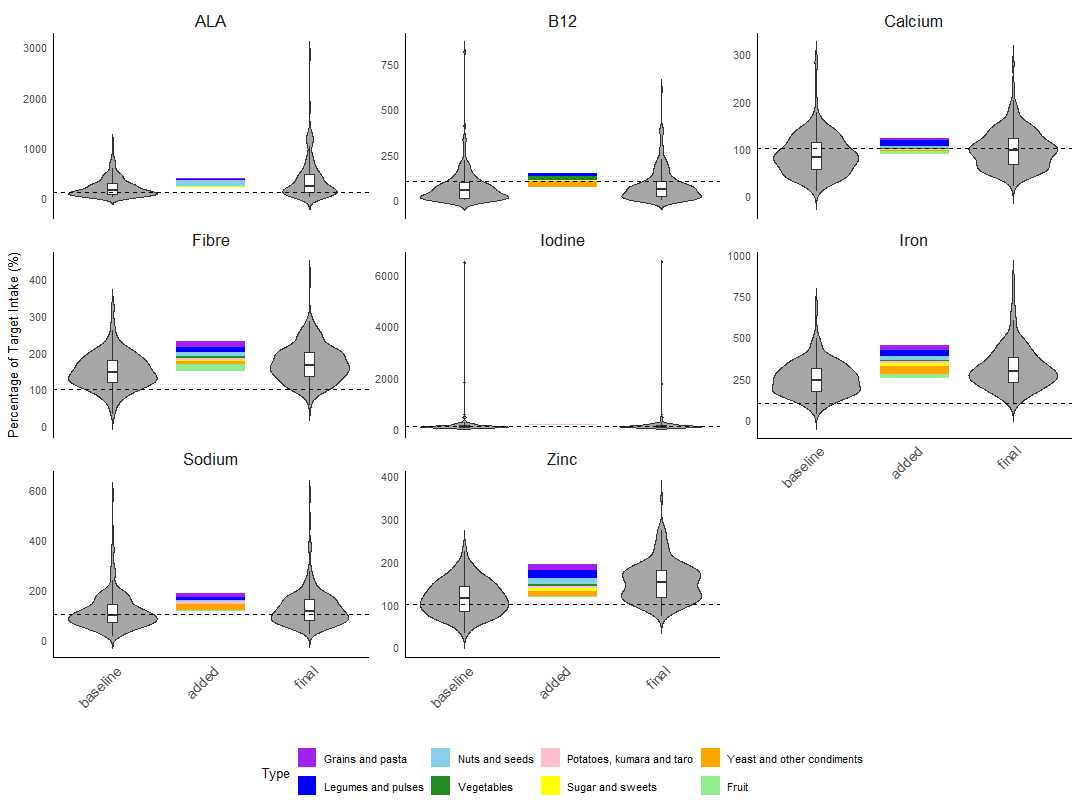

E. **Cluster 2.**

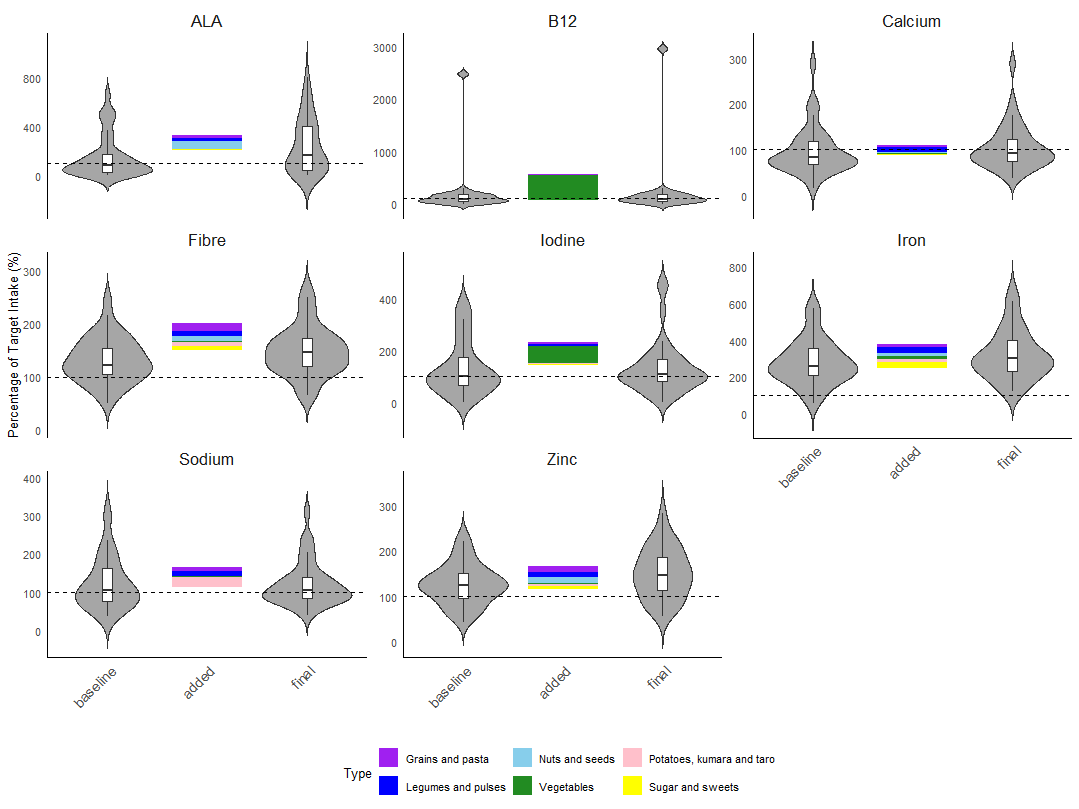

F. **Cluster 3**

**Figure S2.** Comparison of adequacy in dietary fibre, alpha-linolenic acid (ALA) and other micronutrients at baseline and final intakes following diet optimisation in cluster 1 (A), cluster 2 (B) and cluster 3 (C). The dotted line at 100 % indicates the average minimum daily requirement for nutrient. Each violin plot with its box plot represents the overall distribution of energy or nutrient intake, with wider portions in the violin plot representing larger proportion of daily diets at the respective intake quantity (y-axis). The horizontal line in the box plot represents the median of the data with the interquartile ranges at 25^th^ and 75^th^ percentiles (lower and upper edges of the box plot respectively). The line extending from the percentiles represent the range, from the smallest to the largest value within 1.5 times of the IQR. Stacked bar graphs represent the contribution of nutrient as a percentage of the target intake by each food groups. Values representing the nutrient contribution as a percentage of target intake are found in Supplementary Table 1.

**Table S4**. Mean baseline and final intake, and percentage adequacy of other micronutrients across clusters 1 to 3.

|  | Vitamin A (μg) | Vitamin B6 (mg) | Niacin (mg) | Thiamine (mg) | Folate  (μg) | Vitamin C (mg) | Vitamin E (mg) | Potassium (mg) | Magnesium (mg) | Phosphorus (mg) | Selenium (μg) |
| --- | --- | --- | --- | --- | --- | --- | --- | --- | --- | --- | --- |
| Cluster 1 | | | | | | | | | | | |
| Baseline | | | | | | | | | | | |
| Mean intake | 864.32 | 2.09 | 13.4 | 1.47 | 363.4 | 120.2 | 15.6 | 3213.4 | 416.4 | 1083.9 | 39.9 |
| % adequacy | 34.2 | 72.6 | 54.8 | 70.8 | 53.9 | 81.7 | 85.8 | 53.9 | 82.2 | 96.3 | 13.7 |
| Final | | | | | | | | | | | |
| Mean intake | 834.7 | 2.20 | 18.1 | 1.90 | 454.5 | 107.6 | 24.5 | 3551.02 | 592.0 | 1525.6 | 54.2 |
| %adequacy | 32.0 | 83.6 | 84.5 | 89.5 | 72.1 | 78.5 | 94.5 | 71.2 | 98.2 | 100 | 37.9 |
| Cluster 2 | | | | | | | | | | | |
| Baseline | | | | | | | | | | | |
| Mean intake | 781.6 | 1.92 | 15.4 | 1.74 | 384.7 | 118.6 | 16.6 | 3252.8 | 479.7 | 1304.3 | 42.7 |
| % adequacy | 34.4 | 76.9 | 67.2 | 85.1 | 58.5 | 84.6 | 83.1 | 51.8 | 90.8 | 97.4 | 18.5 |
| Final | | | | | | | | | | | |
| Mean intake | 725.3 | 2.05 | 19.4 | 2.07 | 456.7 | 111.7 | 25.1 | 3450.4 | 627.4 | 1677.2 | 54.6 |
| % adequacy | 30.8 | 84.1 | 86.7 | 95.4 | 69.7 | 82.1 | 92.8 | 57.9 | 98.5 | 99.5 | 40.5 |
| Cluster 3 | | | | | | | | | | | |
| Baseline | | | | | | | | | | | |
| Mean intake | 723.1 | 1.86 | 17.4 | 1.60 | 430.4 | 77.6 | 19.4 | 2966.2 | 447.6 | 1394.4 | 53.2 |
| % adequacy | 27.8 | 75.0 | 83.3 | 77.8 | 66.7 | 63.9 | 91.7 | 41.7 | 83.3 | 100 | 22.2 |
| Final | | | | | | | | | | | |
| Mean intake | 727.5 | 1.87 | 21.1 | 1.83 | 459.2 | 76.5 | 26.8 | 3172.9 | 545.5 | 1631.9 | 59.2 |
| % adequacy | 27.8 | 75 | 88.9 | 88.9 | 69.4 | 66.7 | 88.9 | 50.0 | 91.7 | 100 | 47.2 |
